# Principles and test methods of non-contact body thermometry

**DOI:** 10.1101/2022.01.28.22269746

**Authors:** Erik B Beall, Lars Askegaard, Josh Berkesch, Alden C Adolph, Christopher M Hinnerichs, Matthew Schmidt

## Abstract

**Significance:** Far infrared (IR) has a long history in thermometry and febrile screening. Concerns have been raised recently over the accuracy of non-contact body thermometry. Clinical testing with febrile individuals constitutes the standard performance assessment. This is challenging to replicate, which may have inadvertently allowed approval of IR systems that are unable to detect fevers. The ability to test performance without relying on febrile participants would have ramifications for public health, especially if this discovered undisclosed differences in accuracy in widely used devices.

**Aim:** To identify foundational issues in, demonstrate principles of, and develop test methods for non-contact body thermometry.

**Approach:** We review foundational literature and identify confounds impeding performance of IR thermography (IRT) and non-contact IR thermometry (NCIT) for febrile screening and demonstrate corrections for their effects, which would otherwise be unacceptable. Almost none of the devices we are aware of compensate for these confounds. We reverse-engineer surface-to-body temperature relations for several FDA-cleared NCITs. We note their similarity to recently reported bias-to-normal behavior in other devices and determine range of body temperatures for which the device would produce a "normal" (non-febrile) output. Finally, we generate predictable elevated face temperatures in healthy subjects and demonstrate this in several devices.

**Results:** The surface-to-body relationships for two IRT and one NCIT were linear, while all others exhibited nonlinear bias-to-normal behavior that produce normal temperatures when presented with surface temperatures ranging from hypothermia to moderate-to-severe fever. The test method was used in healthy, non-febrile subjects to generate elevated temperatures corresponding to body temperatures from 97.35F to 102.45F. Three out of five systems had negligible sensitivity.

**Conclusions:** This demonstrates an alternative evaluation method without the limitations and risks of febrile patients. These results indicate many devices may be unusable for body thermometry and may be providing a false sense of security for public health surveillance.

## 1 **Introduction**

Core body temperature is used clinically to diagnose conditions associated with abnormally high or low temperatures. Normal body temperature can vary from person to person, by gender, recent activity, food and fluid consumption, mental and physical stress, time of day, and, in people who menstruate, the stage of the menstrual cycle. Clinical studies of oral temperatures acquired with a measurement accuracy of 0.18F (0.1C) in healthy adult humans have found a distribution with a standard deviation of approximately 1F (0.6C) and averages ranging from 97.9F to 98.6F{1,2,3,4}. Oral thermometry, the most widely-used method for vital signs assessment, is sensitive to probe placement, recent speaking or mouth-breathing, recent fluid or food consumption and even air temperature. In-ear thermometry is an oft-used alternative but is affected by ear geometry and debris, often requiring re-measurement attempts. These thermometry methods can be inconvenient due to the time and contact required, and thus, non-contact or other surface-based thermometry methods are increasingly selected.

The physics of long-wavelength IR (LWIR) surface temperature thermometry is well known and covered elsewhere{5, 6}. Briefly however, core body thermometry inferred from a surface temperature of the body relies on availability of a skin region having a local maxima temperature that can be reliably correlated with a core body temperature, good accuracy in measuring that region’s surface temperature, and good accuracy in correcting that surface temperature to an inferred core body temperature. Non-contact body thermometry of a surface uses IR light emitted and reflected by the observed surface and first focuses this light into either a single-pixel sensor (non-contact IR thermometry, or NCIT) or a multi-pixel image sensor (IR thermography, or IRT) and then converts a signal of the intensity of this light into a temperature or an image of temperatures, using a pre-calibrated relation between intensity and temperature, finally applying a pre-calibrated offset between the surface temperature and a reference core body temperature. The choice of single- or multi-pixel is dependent on both how uniform and large the spatial temperature distribution is and on how close to the subject the measurement must be made. A single-pixel NCIT is typically used for forehead, temporal artery or in-ear measurements, while image-based systems are directed towards measurements of the inner canthi made from a greater distance, as per the relevant medical device standards{7,8,9}.

The general operating principle of elevated body temperature detection with IR is as follows: if the core body temperature increases, the appropriate surface temperature will increase nearly proportionally{11}. This general principle is demonstrated in Fig. 1, showing thermal images and thermometry measurements of two human faces having different surface, and therefore core, temperatures. The face on the left belongs to an individual with a normal body temperature, while the face on the right belongs to a febrile individual, as indicated by their oral thermometry readings displayed below each face.

**Fig. 1.**
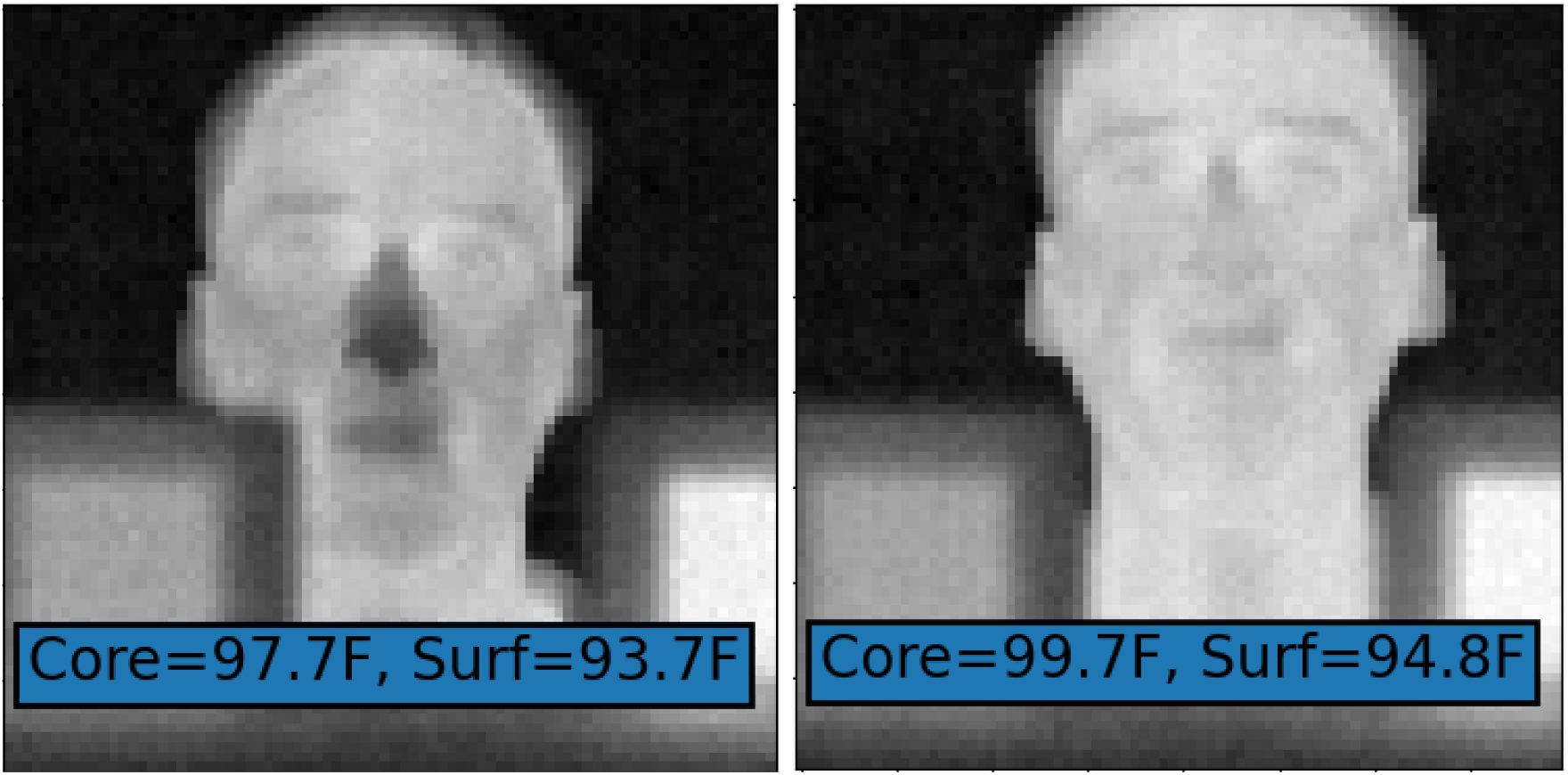
Thermal images of a normal (left) and febrile (right) human face. Inset shows oral (core) and surface thermometry temperatures. Face images shown in this manuscript are of the first author.

This principle is described further, schematically, in Fig. 2 below. This model illustrates warm body heat flowing from core to a cooler environment resulting in a skin temperature that is reduced from the actual core body temperature by the thermal resistance of the skin and air film above it. Because these resistances are approximately constant, if the core temperature is elevated, the skin temperature will also be elevated.

**Fig. 2.**
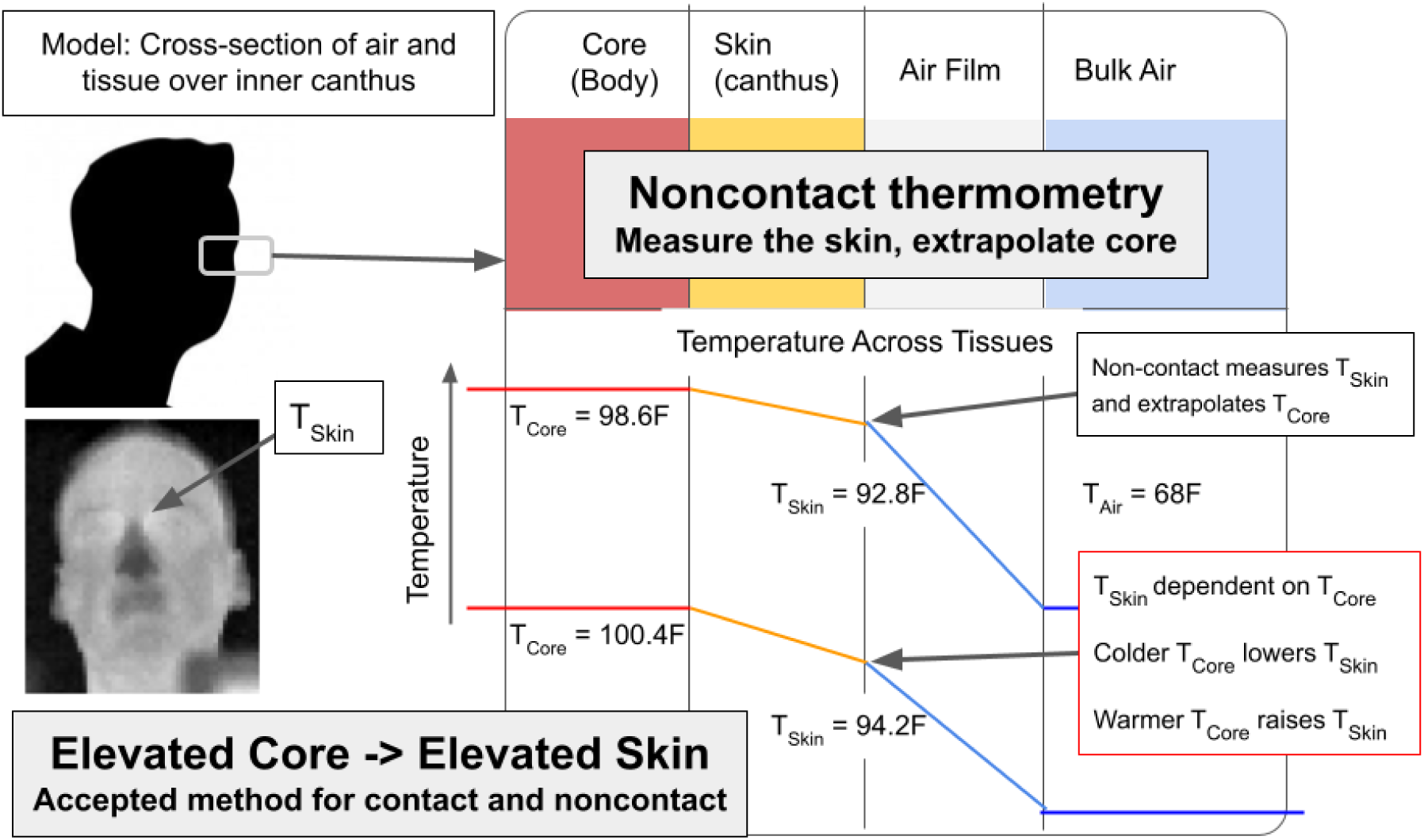
Underlying method of noncontact thermometry illustrated by the upper temperature schematic across components: the measured skin surface temperature is used to infer a corrected core temperature with a relation valid for the air temperature. Underlying method of febrile detection illustrated by the lower temperature schematic: a rise in core temperature produces a rise in skin temperature that allows one to infer the rise in core temperature.

The accuracy of the core body temperature measurement is dependent upon three factors:

1) the accuracy of the underlying calibration relating the pixel signal level to a scene surface temperature,
2) accuracy degradations such as those associated with spatial variations in scene/subject surface temperature that are smaller than the nominal pixel spot size and/or those degradations caused by imperfections in optics and/or focusing, and
3) the correction factor relating a measured surface temperature to a core body thermometry site (e.g. oral).

The first factor, calibration accuracy assessed to a blackbody surface temperature, is well-known and generally solved to the accuracy required for medical purposes for single-pixel sensors, but rarely better than ±2C for multi-pixel image sensors, unless a calibration source is maintained within the scene. There are exceptions for multi-pixel sensors having accuracy below 1C but these are less often used because they are generally an order of magnitude more expensive. Regardless, this seems to be an easy factor to test and confirm with an independent calibration source. However, there are serious limitations to this test, due to the fact that a full-field calibration image is completely insensitive to the inaccuracies caused by the second factor.

The second factor relates to 1) the spatial coverage of a pixel on a surface with varying temperatures and 2) the dependence of a pixel’s signal level on those around it{10}. Regarding spatial coverage, a given pixel maps to an area, or pixel spot, of a surface it is focused upon and the size of this pixel spot is based on the optics, resolution and geometry between the pixel and target. The temperature of skin tends to vary across the surface, and if, for example, the pixel spot is halfway filled with a local maximum and halfway filled with temperatures a degree less than this, then the resulting pixel signal will be about half a degree lower than the maximum. In fact, the actual situation may be somewhat worse than this, due to diffraction and other sources of stray light which is discussed next, but this general concept means there is a maximum distance beyond which the measurement accuracy will degrade. This confound is generally referred to as *distance effect* (DE), and most metrology laboratories have procedures for measuring the magnitude of DE for a given system and application. DE is indirectly treated by the febrile IRT standard by requiring a maximum 1mm nominal pixel spot size{7} but the DE confound is also dependent on focus and optics. In the devices examined in this study, DE is treated by either enforcing a maximum or fixed distance to subject or by applying a distance-based correction and thus is not examined further. NCITs aimed at the forehead or temporal artery from distances greater than a few mm are particularly confounded by this issue, as can be readily demonstrated by placing an NCIT into surface temperature mode, finding a particularly warm location of the forehead by repeat measurements and then acquiring temperatures of this hotspot while increasing distance. The spatial variation of skin temperature across the forehead presents a serious concern that NCITs in real-world usage are unlikely to reliably acquire a local maximum{10} and furthermore, are unlikely to capture said maximum with sufficient single-pixel coverage. While this can be observed with NCITs in surface temperature mode, it is somehow not observed in body temperature mode (possibly due to undocumented normalization of the data, which we discuss later). We are aware of only two NCITs, one of which is examined further in this study, that allow the user to “sweep” the sensor across the forehead for the stated purpose of capturing these maxima, however both of which have been found to bias towards normal body temperature readings{26,27,28,32,33,34,35}.

A second problem arises from the dependence of a given pixel’s signal level on stray light or signal levels corresponding to surrounding pixels. This confound is known in the thermal sensing and imaging field as *size-of-source effect* (SSE){11,12,13,14} but is not assessed in the IRT febrile screening standard{7}. A recent conference publication{10} showed this can cause inner canthi surface temperatures to vary by an order of magnitude more than the surface calibration indicated. The effect can be described as “cooler pixels close enough to warmer pixels will pull the warmer pixels down”, so colder nose, cheeks or forehead regions (or the wearing of a face mask or hat) can cause inner canthi temperatures to read lower by as much as a degree or more, despite this effect being too subtle to visualize from a thermal image. A simple test method to quantify this effect was demonstrated{10, 15}, but to our knowledge, no image-based system currently discloses performance on SSE.

Finally, the surface-to-core temperature correction factor is dependent on physiology and the surface site used to obtain the measurement. This is described as an adjustment in the medical device standard for adjusted-mode body thermometry{8, 9}, and has also been referred to as the physiologic offset and more commonly in industry as the “body mode” offset. To our knowledge, all but one system for body thermometry (out of several dozen we are aware of) use a fixed correction factor that is not dependent on ambient temperature{16}. It is unclear why this is the case, but the relevant medical standards do not discuss any dependence of the correction factor on other confounds, again relying on clinical study to qualify a system as sufficiently accurate, among other issues recently raised regarding the ISO febrile standard{16,17,18}. The strength of an ISO consensus standard{7}, relying on diverse input in its creation, may have led scientists and system designers to assume a fixed offset was indeed sufficient. As shown in the next section, an unacceptably large dependence of the difference between core and canthi or forehead skin temperature on ambient temperature is well-known in the fields of physiology and anesthesiology{19}, as well as in early patent literature for body thermometry{20, 21}, so it is surprising that the concept of a fixed offset has become a de-facto standard. While it is certainly true that if an appropriately narrow operating ambient temperature range were imposed, a fixed offset could indeed provide sufficient accuracy; however, the ambient temperature range allowed by the standard is large enough to induce a 0.64C (1.15F) error in calculated body temperature, which is incompatible with clinical needs because the error is comparable to the lower clinical thresholds for an elevated body temperature (0.5C). The dependence on air temperature used to obtain 0.64C is discussed in Sec 2 and the magnitude of this dependence is shared in Equation 1; it must be pointed out this dependence was determined using a device with minimal SSE and compensated DE and this dependence is likely larger for devices having substantial DE or SSE artifact or other system differences although the factor could in principle be smaller. Nevertheless, the reliance on clinical study{8, 9} for all adjusted-mode thermometry should, in principle, cover the confounds caused by these issues. However, as shown later in this study, in examining a few approved devices which ostensibly adhered to the required clinical testing, we have observed something beyond the above-described accuracy degradation confounds: a bias-to-normal behavior that appears to reduce the actual diagnostic power below the level required for clinical purposes.

To summarize the bias-to-normal performance issue more succinctly, a device that reports a body temperature as being within the normal range of 96.8F to 100.4F regardless of whether the individual being measured has an actual body temperature as low as 95F or as high as 103F is clearly inadequate for clinical purposes (this latter range being referred to as the *normalization range*). This bias-to-normal behavior appears to be present in many non-contact systems, and will allow a device to appear to function normally despite having a poorly calibrated sensor, being used improperly or being operated in adverse operating conditions. Furthermore, the presence of a strong bias-to-normal algorithm can actually appear to give acceptable performance in clinical testing if at least two conditions are met: (1) the ambient conditions are tightly controlled and (2) the febrile portion of the study population consists of febrile temperatures sufficiently outside the normalization range such that they are still detectable as febrile despite the bias-to-normal algorithm. Simply, a bias-to-normal algorithm is an algorithm that produces apparent, not actual, diagnostic performance of a device.

### 1.1 Foundational Literature - Physiology

We turn now to the physiology underlying the relationship between human skin and core temperatures, because this is essential to obtaining an accurate core body temperature with non-contact methods in real-world scenarios. The physiologic offset is the reduction in temperature between core body temperature and a well-chosen patch of skin. For the inner canthus, which has minimal thickness and vasomotion of tissue over the underlying primary artery, the slope of the relationship is driven by the thermal resistivity and the environmental conditions. To first order approximation, the total heat flux from core blood in the angular artery through this area of skin to the environment is set by the thermal resistivity of the skin, the conductivity and convection of mostly still air and the radiation balance to the surrounding environment. Extensive modeling of heat flow from core to skin has evolved since the original Pennes bio-heat equation{22,23,24}, but in the human inner canthi, the effect is approximated well by a linear or low-order polynomial relationship, which has been documented in the literature over typical ambient air temperatures between 30F and 100F{19}. See for example Figures 10 and 11 in this{21} 2003 US patent. (For context, further below we reference Section 2.1, which introduces Equation 1 to describe the effect of ambient temperature on the skin-to-core physiologic offset and Equation 2 to infer a core temperature from surface and ambient temperatures.)

Understanding the relationship between skin and core temperature is essential to monitoring core temperature, in particular for monitoring hypothermia during anesthesiology, which has been in active study for over 20 years. Ikeda et al{19} examined the impact of ambient air temperature on the offset between forehead skin and core temperature. They found a 20% dependence on ambient temperature, stating “Transferring a patient from a typical 20C operating room to a typical 25C postanesthesia care area, however, would reduce the core-to-forehead difference approximately 1C.”{19}.

The same research group, led by Dr Daniel Sessler, later performed a series of challenging and carefully run clinical studies of the monitoring and regulation of temperature, and made a series of valuable observations that are relevant and we collect them here. First, the essential performance required is ±0.5C for anesthesia purposes, because a 1C change has distinct physiologic consequences{25}. In further studies, the Sessler group examined several external thermometry methods (e.g. non-contact as well as contact) and compared accuracy against direct thermometry at various internal sites, such as the tympanic membrane{26,27,28}. It is important to point out that these anesthesiology studies that mention “tympanic” measurements use true tympanic measurements, which are highly challenging to perform as they involve directly contacting the tympanic membrane with a thermocouple and verifying actual contact, while studies outside of anesthesiology will refer to so-called “tympanic” measurements that are actually performed with aural NCIT thermometers that typically cannot see the tympanic membrane. In the field of thermography, cavity radiation is a useful tool to obtain accurate measurements, so the argument that an aural NCIT placed into a somewhat tortuous cavity enclosed at one end by the tympanic membrane could still accurately track true tympanic temperature is attractive (but see Daanen{29} for problems with aural geometry). However, these in-ear NCITs (as well as the temporal artery NCITs) have been shown by the same group to be significantly different from true tympanic temperatures, and unreliable for anesthesia monitoring purposes: “As normally used, that is directed into the aural canal or near the temporal artery, infrared systems are insufficiently accurate for clinical use … In light of their poor performance, it seems unfortunate that they have become so popular.”{25}. We mention this distinction because it is a real, yet widely unrecognized, difference between true tympanic measurement and the vastly more common aural NCIT so-called tympanic measurement. The widespread use of different non-core body sites as the “gold standard” is problematic when the terminology is inconsistent because it can lead to biased results and makes interpretation more difficult.

One such Sessler group study examined aural NCIT, oral thermometry, “deep forehead”, and a temporal artery (TA) NCIT against bladder core body thermometry{26}. The Bland-Altman plots (figure 3 from that study{26} shows TA minus bladder vs bladder) show clear bias-to-normal behavior for the TA NCIT, in particular having a negative slope, meaning a given range of actual core temperatures may be reported as lying within a much smaller range. All NCIT methods were insufficient, particularly the temporal artery NCIT, followed by aural NCIT then deep forehead, with only oral thermometry being acceptable for clinical purposes. Their focus on accurately monitoring core body temperature is driven by more than scientific curiosity: “Larach et al. estimated that the risk of dying after a malignant hyperthermia crises [sic] was an order of magnitude higher in unmonitored patients, presumably because temperature monitoring facilitated speedy diagnosis.”{30, 31}. *Thus, at the risk of stating the obvious, there is a clear clinical mandate for maintaining accuracy to within ±0.5C*. If NCITs are inaccurately estimating core body temperatures, it could have vital implications for quality in clinical care, because many practices rely on some form of NCIT for quick and easy body temperature measurement during vital signs assessment.

**Fig. 3.**
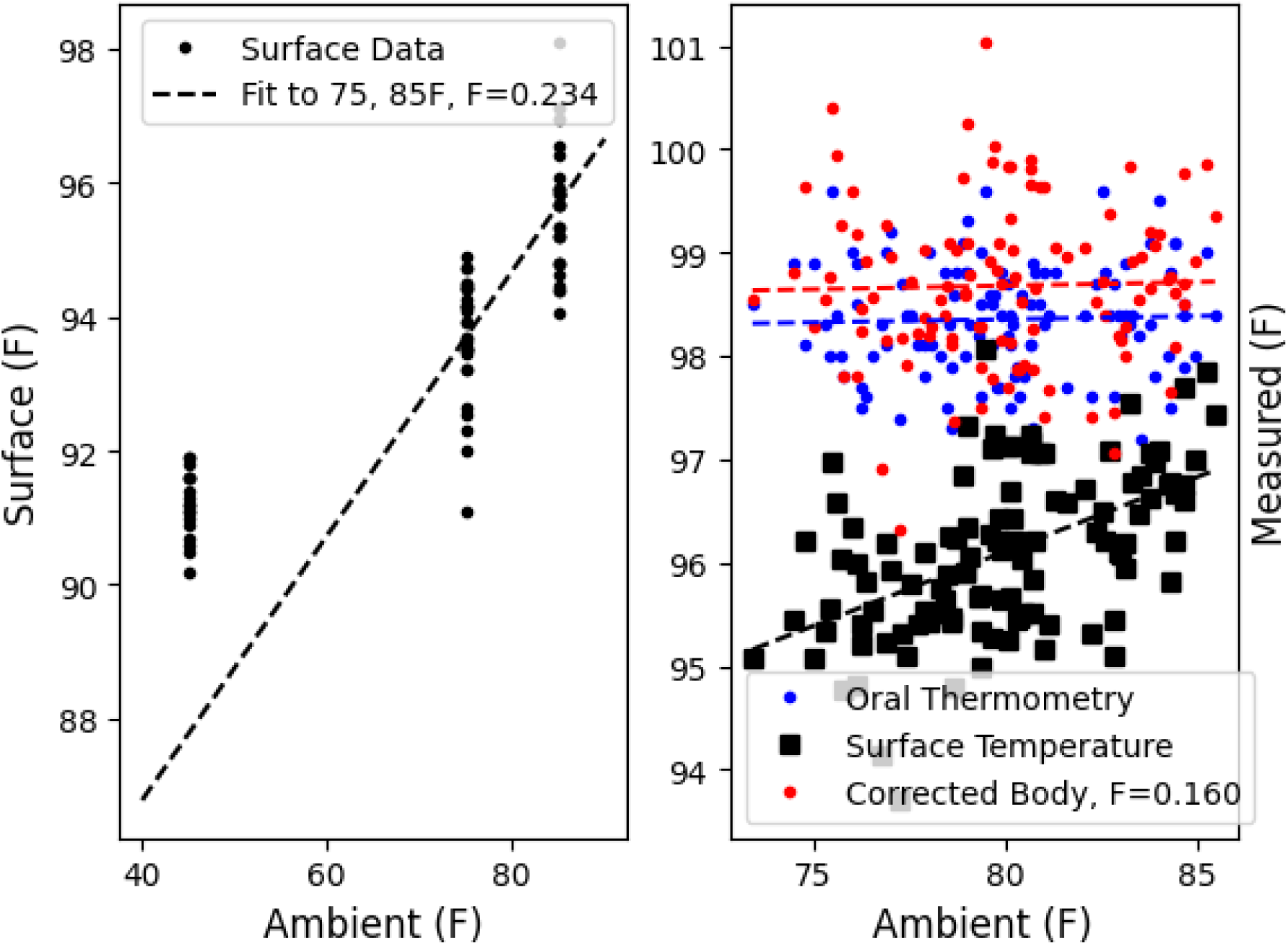
**a)** Inner canthi skin surface temperatures at three ambient temperatures in a pilot study of 32 individuals. The spread in skin temperatures was confounded by improperly recorded air temperatures and an approximately 1C SSE confound that was not recognized at the time of the pilot study, while the 45F data collection was confounded by an equilibration time of under 2 minutes and therefore ignored in the fit. **b)** Data recorded in the present study showing inner canthi skin surface temperature in black squares, oral thermometry in blue and corrected body thermometry in red versus effective environmental temperature.

Several other groups have studied NCIT accuracy with similar findings of poor accuracy and bias-to-normal behavior{32,33,34}. In one particular study, Low et al{32} investigated the same TA NCIT used in the Langham study with a wireless ingestible core body thermometer and active manipulation of core body temperature and found similar behavior, specifically that the Exergen TA NCIT measurements were decoupled from actual core body temperature increases. Intriguingly, the authors examined US Patent 6292685{20} covering this device and reproduced the central equation that bears the same general form as found in other patent literature{21} and in a pilot study{35} we describe in Section 2.1. This equation has a linear dependence on ambient temperature, with the slope based on perfusion and specific heat capacity of blood.

The bottom line is that groups with expertise in physiology have shown that the ambient dependence of the physiologic offset is large enough that it must be accounted for to obtain a sufficiently accurate surface-to-core correction, which fortunately is easy to do with an appropriately careful air temperature measurement.

### 1.2 Foundational Literature - Non-contact Fever Screening using IRT systems

We turn now to the historical development of non-contact thermographic body temperature screening systems and the relevant international standard{7}. Studies of IRT for febrile screening lie in three groups: 1) post-hoc febrile discrimination (poor generalizability), 2) prospective discrimination and secondary screening of IRT-positive subjects (modest generalizability), and 3) prospective discrimination with secondary screening of all subjects (best generalizability). Post-hoc febrile discrimination refers to the practice of acquiring skin temperatures, deriving a correction or threshold from this same data that is then applied to this same data before deriving sensitivity and specificity (we will refer to this type of methodological error as a “post-hoc threshold”). This practice means the threshold may not apply to data collected by other groups, to data collected with different equipment (even of the same model) or even to the same group using the same equipment but on different days. Due to the dependence of IRT core body measurement on ambient temperature, SSE, DE, and other confounds, any post-hoc analysis like this is unlikely to generalize to later work, unless these confounds are all accounted for both in the threshold-setting study and all subsequent operations. Given two important confounds (SSE and DE) that can sum to several degrees Celsius were not published as relevant confounds for body thermometry until 2021, as well as the dependence of of the skin-to-core offset on ambient temperature, it is unclear how to interpret past studies relying on post-hoc threshold-determination{36,37,38}. While there is a considerable body of work addressing the SSE, it has apparently escaped notice by the developers, and most users, of the body thermometry standards.

A smaller list of studies did not rely on a post-hoc threshold. An 11-month study from 2005 of Dengue screening with IRT at Taiwan’s two main airports acquired over 8M passengers, with 22,000 identified as febrile by IRT{40} (IRT methods were not described). Of these, 3,011 were confirmed by aural NCIT and by serological analysis, 40 were identified as carrying Dengue, with 33 being actively viremic. The authors of that study used the statistics of the nationwide Dengue screening program to infer that these two screening stations captured the majority of Dengue infections traveling into Taiwan.

A particularly influential study was that of Ring et al.{41}, performed in a pediatric hospital in Poland in 2008 with 191 subjects between the ages of 1 and 18 yrs, with axilla contact thermometry and aural NCIT performed in parallel with IRT. Because this study was performed with deep knowledge of the ISO and IEC writing group’s early drafts defining the minimum standards needed for the use of a dedicated screening thermograph, it helped define{42} the underlying science and method in the final febrile screening standard{7}, using the data collected in this study and the impact of proper positioning of the head shown by the Glamorgan Protocol{43}.

The study of Nguyen et al. 2008{44} collected IRT, oral thermometry and self-reported fever in patients arriving at the emergency departments of hospitals in three major US cities. Out of 2873 patients, 476 self-reported fever (16.6%) but only 64 had oral-confirmed fever (2.2%). *Thus, self-report, even in an emergency department, was found to be a poor predictor of febrility (sensitivity of 75% but PPV of only 10.1%)*. IRT performance varied across equipment, with only two of the three IRTs producing better sensitivity than self-report. OptoTherm and Device #5 were 91 and 90% sensitive and 86.0 and 80.0% specific, respectively, with r-values of 0.43 (Optotherm), 0.42 (Device #5), but only 0.14 for the third (Wahl).

In Zhou et al.{45}, the authors obtained IRT and 3-minute oral thermometry in 596 subjects, 47 of whom were febrile (>37.5C via oral) and generated 17 facial locations using hand-drawn regions-of-interest (ROI) for each subject. Receiver operating characteristic (ROC) analysis for each ROI provided the best sensitivities in three regions: 0.95 for inner canthi, 0.97 for full face and 0.865 for full forehead. The correlations for both inner canthi and full face ROIs were close to 0.8. Zhou et al. also made the point that low-grade fever would be the most likely target for screening, since mild and moderate febrile subjects are less likely to modify behaviors.

We are aware of only one prospective threshold study to have acquired viral status of both IRT-positive and IRT-negative subjects{46}, relying on an outbreak of Swine flu aboard a United States Navy vessel in the Pacific Ocean in 2011 by a co-author on the present paper, Dr Hinnerichs. In Hinnerichs et al., the threshold was set daily with a core-to-skin calibration on 10 afebrile individuals at the start of each day and the calibrated IRT and oral thermometry were collected in 320 subjects, 84 of whom were later confirmed positive for swine flu by laboratory PCR. The IRT operator was blinded as to the febrility status of each individual (and PCR results were not provided until after the IRT collection was complete). The environment was maintained between 71 and 73F during the data collection. The study achieved r-values of 0.90 and 0.87 (male and female respectively) and sensitivity of 84.5% with specificity of 97.5%. ROC analysis showed the ability to discriminate febrile from nonfebrile 91% of the time. The study further tested for any gender difference in sensitivity and found none, albeit in a relatively homogeneous population consisting predominantly of active service members.

Finally, due to the COVID-19 pandemic, the febrile screening market saw an explosion of supply of “automated” IRT systems from new vendors, many of whom were marketing low-resolution thermopile IRT systems that did not have an IR calibration source to obtain the required accuracy. A recent study (involving a co-author of this study, Dr Hinnerichs) examined the performance of seven recently-introduced automated IRT products and found all exhibited substantial bias-to-normal behavior, meaning their outputs “systematically biased to the mean human temperature”{47}. This publication was, to our knowledge, the first to identify and explicitly state the presence of a systematic bias that could produce harm to public health, and raised the point that no known physics or physiologic model could justify the use of a biasing algorithm with the normalization ranges as wide as those observed. The authors noted one limitation of the study was their inability to incorporate human subjects, due to the unavailability and safety of working with febrile (infectious) individuals. The authors pointed out their study should translate directly to real human faces, but as the work on SSE has shown{16}, it is likely the systems investigated would perform even worse on real human subject faces.

### 1.3 Medical Device Standards for Non-Contact Thermometry

The ISO standard for medical thermometry, IEC 80601-2-56:2017{8} (hereafter referred to as the “thermometry standard”), describes devices that fall into two primary categories: direct and adjusted. Direct relies on the sensor reaching equilibrium with the body site (e.g. oral, rectal, axillary) while adjusted takes one or more sensor readings and adjusts to a surrogate for a body site. Direct mode devices may be certified via laboratory testing, while adjusted mode devices must also undergo clinical testing for accuracy in human subjects against a direct-mode reference thermometer at a specified reference site (e.g. oral). NCIT and IRT both belong to adjusted-mode thermometry, although at time of writing, no IRT device has undergone testing to the thermometry standard. Note, we are aware of one such device that might be considered an IRT (relying on a multi-pixel sensor rather than a single-pixel) used for NCIT, the WelloStationX, but it is unclear whether this is the case given the sparse documentation available at this time. Rather, there is a separate standard describing the performance requirements for IRT devices intended solely for febrile screening, IEC 80601-2-59:2017{7} (hereafter referred to as the “screening standard”), which does not require clinical testing. The screening standard describes laboratory measurements for an IRT system comprised of either an IR camera with sufficient absolute accuracy or an IR camera paired with an IR calibration source (blackbody) to achieve a total system uncertainty of ±0.5C. Recent publications have pointed out shortcomings in the standard and proposed enhancements or modifications{16,17,18}. As discussed earlier in the introduction and results from our pilot study{36}, the lack of an ambient-dependent adjustment is a major confound for both IRT and NCIT devices. Further, the lack of a specification for the pixel independence or SSE{16} is a major problem for the reliability of IRT and NCIT devices.

We could end this section by drawing attention to the use of clinical testing for any adjusted-mode thermometry and urge the same clinical testing be required for the screening standard as well. Clinical testing should be used because it can provide validation of device performance while being operated as intended. However, devices that appear to exhibit severe bias-to-normal behavior have been cleared for marketing by the FDA as clinical electronic thermometers (product code FLL). The results in Section 2 indicate the reporting to the FDA of clinical tests performed on some devices may have been inadequate. Given the expertise and care required to perform clinical testing is beyond the reach of many manufacturers, it may be helpful to require clinical testing be performed by independent entities, but the testing currently specified is too challenging to be applied by independent test laboratories due to the reliance on convenience samples of febrility that may pose too much risk to be used outside of clinical environments. New test methods that avoid the need for febrile subjects could ameliorate these challenges.

## 2 Rationale for the Test Methods

### 2.1 Dependence of the Skin Surface Temperature on Core and Ambient Temperatures

The dependence of inner canthi skin temperature on core and ambient temperatures was briefly described in Sec. 1.1 with references to anesthesiology and physiology literature and US patent literature. The approximately linear form of this dependence has been confirmed by one of the authors in a pilot study incorporating 32 volunteers performed in June 2020{36}, where each individual’s facial maxima was recorded while the subject had been equilibrated at three stable ambient temperatures ranging from 45F to 85F (the 45F data was excluded due to incomplete equilibration) and this data compared with oral thermometry. This limited pilot study found a 24% dependence of the offset between skin and core (oral) temperature on ambient temperature, as shown in Fig. 3a.

A static analysis of the heat flow across skin (described in Sec. 2.3) results in conversions between inner canthi skin and core (oral as the surrogate) shown in the following equations:

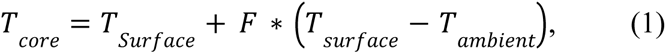

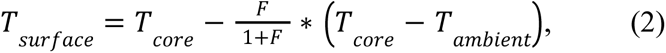

where T_core_ is the core (e.g. oral) body temperature, T_surface_ is the selected skin surface temperature, T_ambient_ is the environmental temperature, and F is the dependence of the physiologic offset on ambient temperature. The physiologic factor F is dependent on skin site and is expected to have some level of variability from person-to-person. For a particular IRT or NCIT, F may also incorporate some level of confound from SSE and DE and other equipment-specific characteristics, so it must be pointed out this factor is likely device-dependent but we expect it to be a fixed factor for a given device model over a reasonable ambient temperature range. In the pilot study described above, the slope of the fit in Fig. 3a resulted in F = 0.24. This has been revised to **F = 0.1595** using the datasets collected in this study and described in Sec 3.1, all of which were collected after the implementation of a SSE correction{10} as shown in Fig. 3b, which shows inner canthi surface measurements, oral thermometry and corrected body temperatures, with the body correction derived from the linear fit of the offset between oral thermometry and surface temperature to the environmental temperature. Note the correction does not include the intercept from the linear fit so the resulting corrected body estimate is shifted slightly higher than the oral thermometry values from which the fit was derived. This factor is specific to the inner canthus, and other body sites have different factors, which is driven by the amount of insulation between fully perfused tissue and the environment. For example, an aural NCIT inspected in the next section (Fig. 4) appears to use a lower factor of F = 0.096 (the fitted slope minus 1), while the forehead site has slightly higher insulation and thus would use a higher value for F than the inner canthus (note, the forehead F-value was not measured here).

**Fig. 4.**
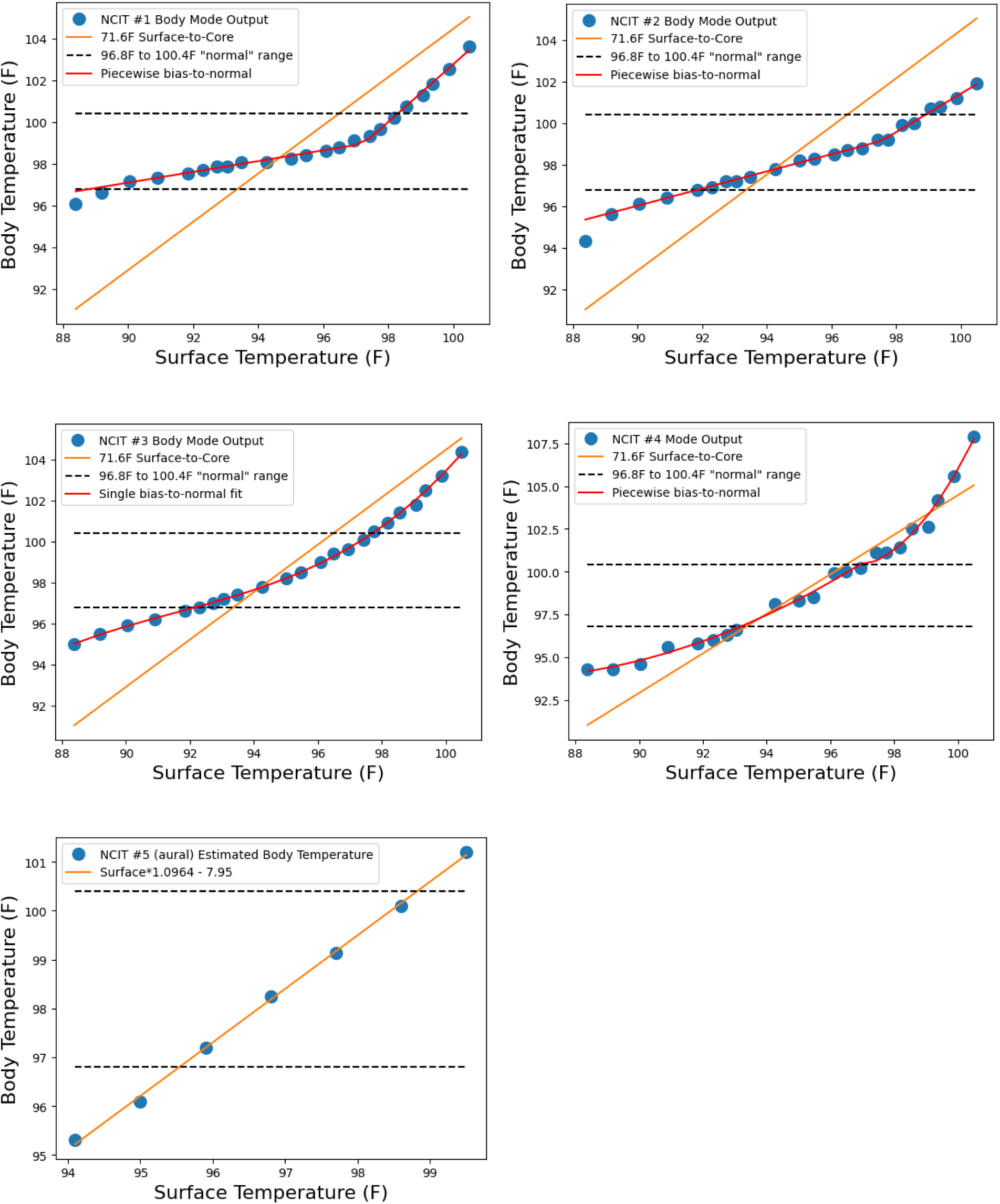
Surface-to-Core transfer curves for 5 NCIT devices denoted in Table 1.

**Table 1.**
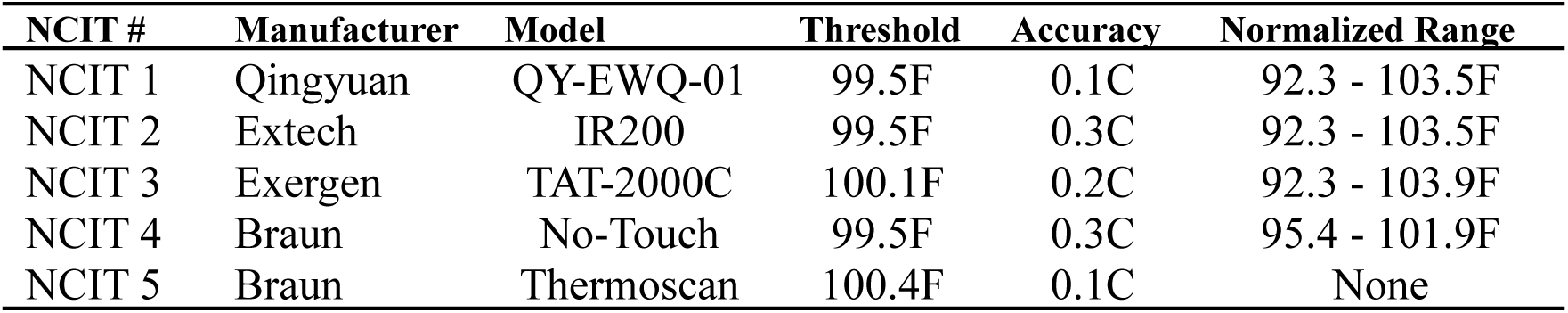
Specifications of the five NCIT devices inspected for transfer curves.

| NCIT # | Manufacturer | Model | Threshold | Accuracy | Normalized Range |
| --- | --- | --- | --- | --- | --- |
| NCIT 1 | Qingyuan | QY-EWQ-01 | 99.5F | 0.1C | 92.3 - 103.5F |
| NCIT 2 | Extech | IR200 | 99.5F | 0.3C | 92.3 - 103.5F |
| NCIT 3 | Exergen | TAT-2000C | 100.1F | 0.2C | 92.3 - 103.9F |
| NCIT 4 | Braun | No-Touch | 99.5F | 0.3C | 95.4 - 101.9F |
| NCIT 5 | Braun | Thermoscan | 100.4F | 0.1C | None |

### 2.2 Observed Bias-to-Normal Algorithms

Some NCIT devices can be alternated between surface temperature and body temperature modes. This makes it straightforward to extract the surface-to-core transfer curve by directing a device at a calibration source and alternating between modes, while sweeping the calibration source through the expected surface temperature range in suitable steps. For those devices that do not have a surface temperature mode, a limited form of this process can be performed with body mode only in order to inspect the linearity, if not the exact numeric offset, of the surface-to-core curve. These can also be performed for some IRT devices, but not those that require the presence of a face in order to function. For this reason, we only investigated these transfer curves in NCITs. Once sufficient data is collected, an approximation of the internal algorithm can be reverse-engineered by piecewise fitting to arrive at a suitably close approximation.

The four forehead NCITs and one aural NCIT in Table 1 were examined for surface-to-core transfer functions (Note NCIT #3 is also represented in Table 2 as Device #3):. The first four devices exhibited strong bias-to-normal behavior, where the highest body temperature that would be biased into the normal range (outputting a body temperature below the device’s febrile threshold) is given in the last column of Table 1. The first three NCITs did not produce a temperature below mild hypothermia (95F, or 35C) for the lowest simulated body temperature used which was 92.34F. The aural NCIT and Device #5 and Device #1 (see Table 2) IRTs did not exhibit any bias-to-normal behavior. The Device #2 and Device #4 IRTs were not assessed in this manner. The aural NCIT (NCIT #5) uses a smaller physiologic offset (the factor in Eqn. 1 would be 5% for aural canal, as compared to the 20-25% used in Device #1 for inner canthi) due to the aural site being closer to core body temperature than the forehead which also has the consequence of utilizing a narrower range of output values. None of the NCIT devices had any apparent dependence of the physiologic offset on ambient temperature, while Device #1 is known to use Equation 2 shown as the 71.6F Surface-to-Core line and which shifts the transfer curve up and down depending on the ambient temperature.

**Table 2.** Specifications of the four IRT and NCIT devices used in the study. See Table S5 for more detailed specifications.

| Device # | Model | Type | Threshold | Accuracy | Acquisition time | Output |
| --- | --- | --- | --- | --- | --- | --- |
| Device 1 | FIP | IRT | 100.4F | 0.3C | 2sec | Body |
| Device 2 | DT982Y | IRT | 99.14F | 0.5C | 0.5sec | Body |
| Device 3 | TAT-2000C | NCIT | 100.1F | 0.2C | 5sec | Body |
| Device 4 | Seek Scan | IRT | 100.4F | 0.3C | 3-30sec | Body |
| Device 5 | T540-EST-42 | IRT | **+1.8F** | 0.3C | 0.5sec | Surface |

An immediate need is a clinical test method capable of generating arbitrary realistic febrile faces, without requiring the risk or restrictions associated with using actual febrile subjects. The motivation for improving testing is to improve how the devices perform in the population they are most likely to be used within. The most severe fevers will either be reduced by antipyretics below the severe range or will remain out of the general population because they are aware they have a fever. Additionally, febrile screening that only performs at 103F and above while biasing temperatures between 100 and 103F to normal is actively harmful. Therefore, the clinical testing requirement must attempt to specify a more uniform range of febrile subjects (e.g. weighted more equally between mild, moderate and severe). The central challenges are that the availability of a useful range of febrile subjects cannot be guaranteed, and proximity to febrile subjects is associated with unacceptable risk.

### 2.3 A Test Method to Simulate Feverish Individuals

As discussed above, non-contact medical thermometry is based on a linkage between core temperature and skin temperature. When core temperature rises, skin temperature rises, and this offset between skin and core can be found using physiologic models of the flow of heat to the environment via the skin. To good approximation, the same result can be found with simple static thermal analysis (Fig. 5), by approximating the tissue above a near-surface primary artery as a passive thermal insulator. The flux of heat flowing across this insulator is dependent on the environmental insulation and temperature. Assuming fairly still air and a radiative temperature close to the air temperature, the flux is proportional to the difference in temperature divided by the total thermal resistance. This leads to Equations 1 and 2 in Section 1.1, which is calibrated for human inner canthi using one thermal camera system. It should be pointed out that the ambient dependence factor may differ due to subtle dependencies of the non-contact system used, including wavelength dependencies and image sensor spot size on target.

**Fig. 5.**
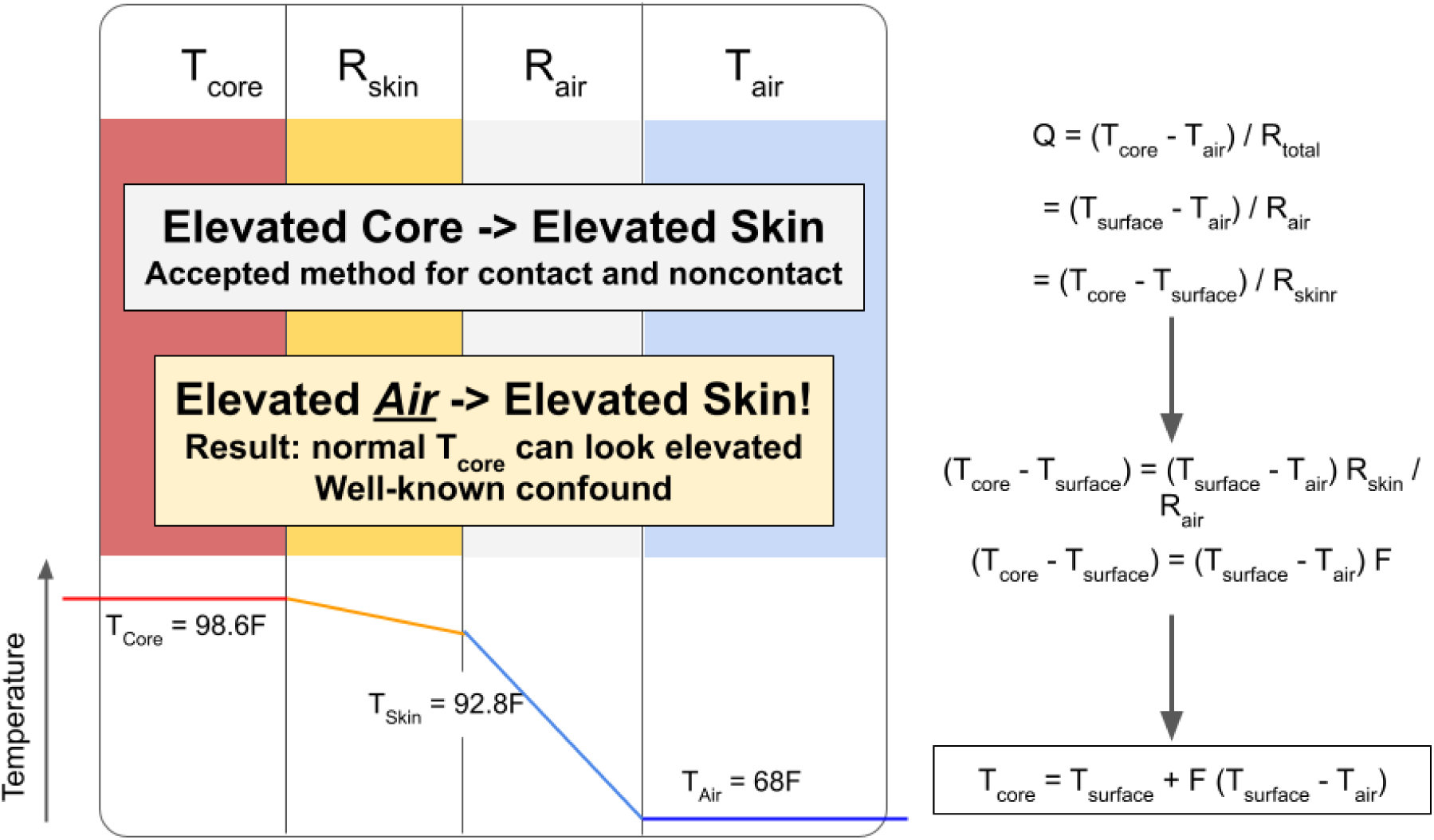
Static thermal analysis of inner canthus. Note the lower inset which is contrasted with that from Fig. 2 above it, elevated air temperature causes elevated skin temperature in a manner that is measurably equivalent to elevated core temperature. This is a well-known confound that is taken advantage of in this study to devise a test of device performance without requiring febrile subjects.

This can be extended to the dynamic evolution of skin temperatures as the environmental temperature is changed. Fig. 6 shows Device #1 operated in body mode on a subject equilibrated to one room temperature (first column, 73F air temperature, 93.4F skin temperature and 98.2F calculated body temperature) and then moving to a second, elevated air temperature (86.6 to 87.3F air temperature, columns 2 through 4, showing time after first image). Initially, the skin temperature matches the previous skin temperature but rises rapidly, then slows and stabilizes. The skin temperature rises by over 3F during equilibration to stabilize at a value that produces a realistic calculated core body temperature, relative to that in the first panel, while the body temperature is underestimated when measured before equilibration has nearly finished.

**Fig. 6.**
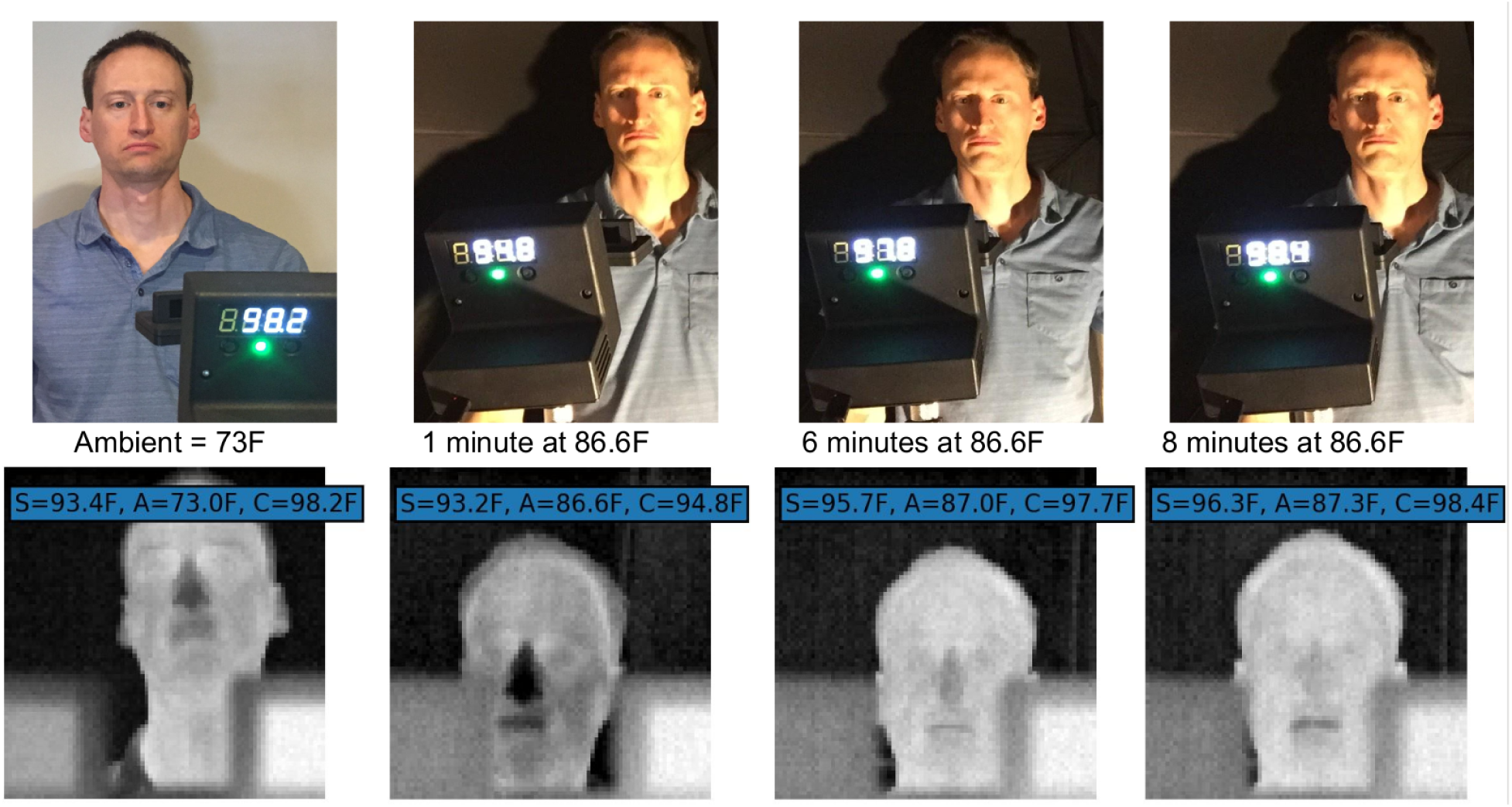
Exposure to different ambient (A) temperatures adjusts the surface (S) temperature, but may not affect core (C) temperatures. Example of an equilibrated subject moving from one environment (73F) to a warmer environment (86.6F) and the gradual evolution of the surface temperature. Calculation of core temperature based on Eq 1 (using F=0.234 from the pilot study) is correct only when the subject has equilibrated to the environment, as shown in the first and last panels only, with errors shown in the intervening panels.

This relationship (Eq. 1) combined with the underlying principle of non-contact thermometry suggests a method for generating test data in human subjects that would correspond to a nearly arbitrary range of actual core temperatures, while relying only on subjects with a normal range of core temperatures. In this method, subjects’ skin is allowed to equilibrate to a first environment with altered air temperature selected using Equations 1 and 2, and the simulated ***elevated body temperatures*** determined from the oral thermometry, Equations 1 and 2, and the elevated air temperature (or, if surface temperature is measured, this can be calculated using Equation 1). Next, the test equipment is operated from within a second controlled environment having a view into a protected portion of the first environment. The ***detected body temperatures*** are recorded from this second cooler environment upon confirmation of equilibration, by briefly opening a pathway to the subject’s skin, while carefully controlling to reduce equilibration to this second environment from air moving from the second to first environment. The devices can then be assessed for sensitivity, specificity and any other metric desired by comparing the elevated and detected body temperatures.

The equilibration process follows an approximately exponential decay between the previous environmental temperature to the new environmental temperature with a characteristic time constant. Fig. 7 shows a sequence of inner canthus skin temperature measurements after exposure to blown, heated air, which is not to be considered a suitable simulation of equilibration but used solely for illustration of time constants. For this test method, a minimum equilibration time of 10 minutes was set based on preliminary study confirming sufficient equilibration within several time constants. More work is planned to further elucidate the physiology of equilibration.

**Fig. 7.**
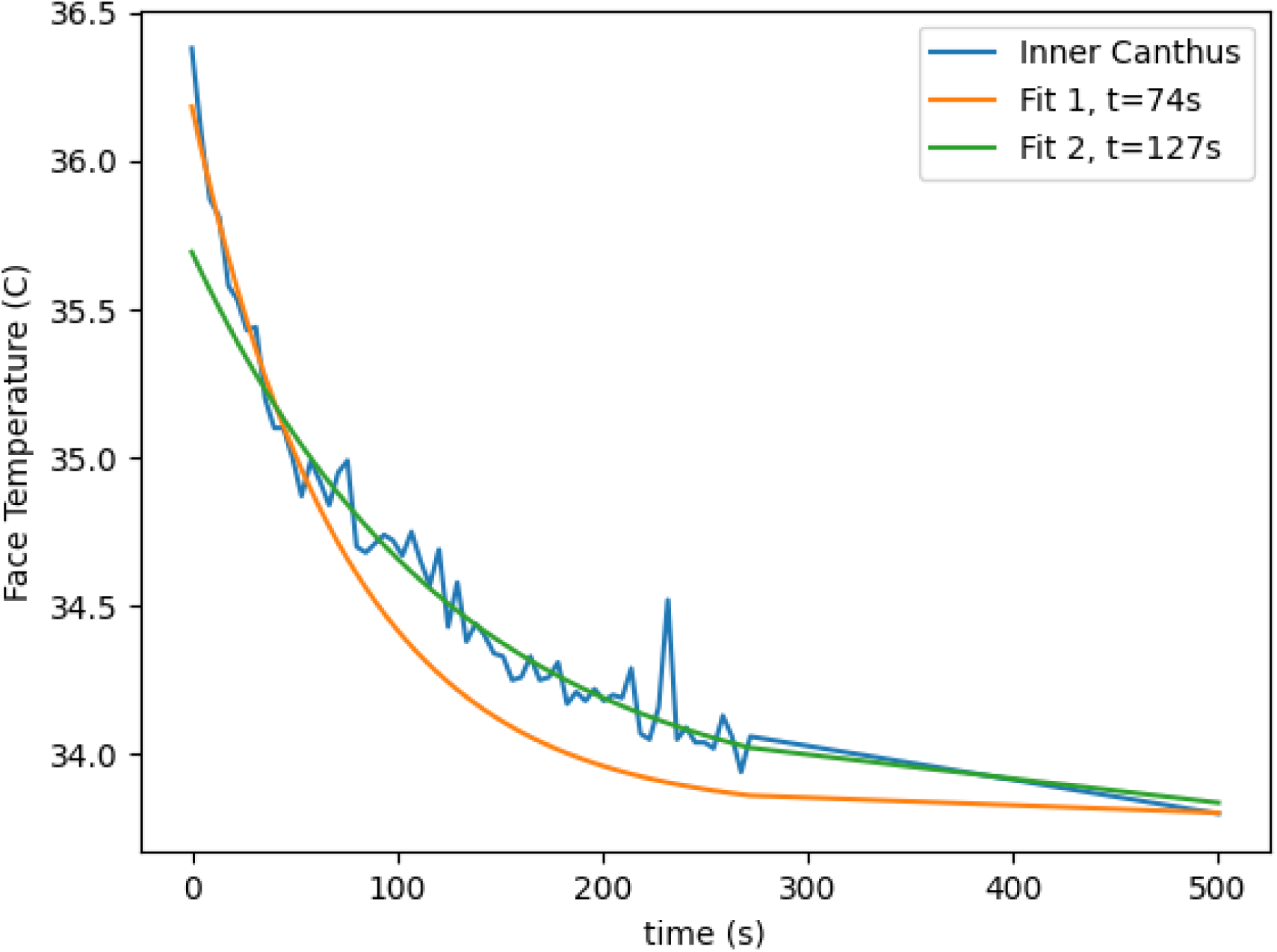
Equilibration curve for skin exposed to a forced warm air for 60 seconds and then ambient air immediately prior to the start of acquisition. Fitting the exponential decay limited to short and longer time bounds found a time constant of 74 seconds for a faster component and 127 seconds for a slower component. Note, this synthetic scenario is not intended to replicate equilibration to moderately still air, only to illustrate the concept.

**Fig. 8.**
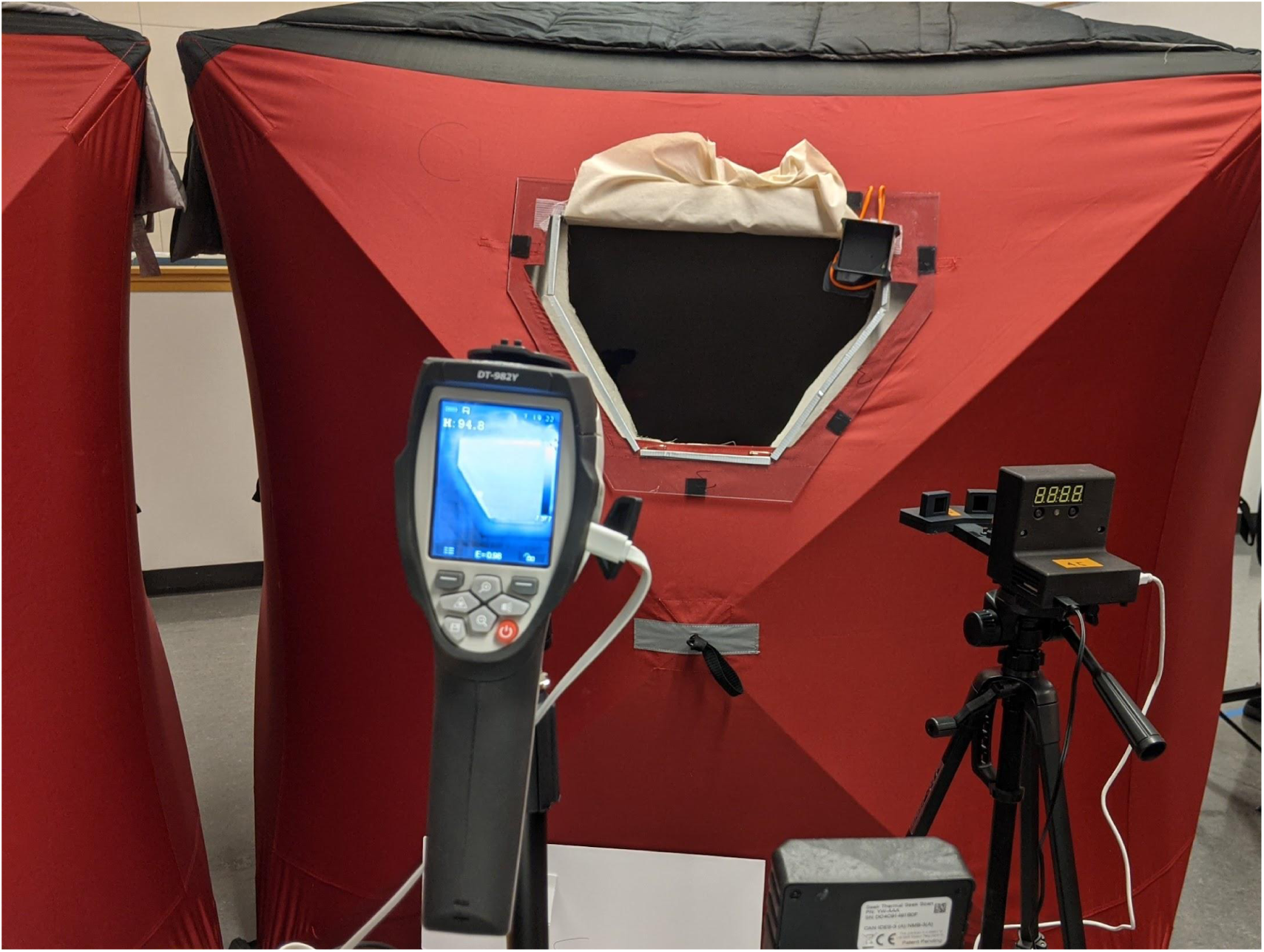
Environment (tent) with opening cover removed and radiative shield lifted and tucked above the opening frame where each subject would stand for Snapshot measurements after completion of 10 minutes of Equilibration measurements. Device #4’s blackbody can be seen at the top right of the frame, Device #1 is shown to the right on a tripod stand, Device #2 is shown to the left. Not visible in this image: Device #3 and Device #5.

Several caveats are important to mention. First, the equilibration proceeds via conductive and radiative pathways, with more than half of the heat flow via radiation, so if the first environment’s radiative background temperature is significantly different from its air temperature, the equilibration will be to a different *effective* temperature. In many situations the background heat-carrying infrared radiation is coming from surfaces that are within a few degrees of the air temperature and as a result the effective temperature is within a degree of the air temperature. However this cannot be guaranteed in general and must be checked (see the *Supplement - Radiative and Effective Temperature*). This can be accounted for by taking an appropriately weighted sum of the radiative and air temperatures (e.g. 70% of radiative plus 30% of air) but it is easier in practice to use radiative shielding, such as sheets hung from a frame, suspended in air such that they match the air temperature. This can be verified during testing using an NCIT in surface temperature mode to verify radiative surfaces are within a suitable range of the air temperature, e.g. within 0.6C should produce an error no greater than 0.1C in calculated body temperature. Alternatively, an IRT used for tracking equilibration can serve the same purpose. Secondly, drafts will affect equilibration by reducing the boundary air layer at the skin so this must be minimized. Thirdly, if the temperature is not stable in both environments, the equilibration will not be stable, so the stability of the air temperature must also be monitored in both environments. If the radiative environment is uncoupled from the air temperature due to convection or too wide of a control window, the background radiance and air temperatures can be used to obtain an accurate estimate via a combined, effective temperature. Fourth, once equilibration is reached, the subject’s skin must be exposed to the test equipment, and this must be done without or before significant change in the subject’s skin due to exposure to the test equipment environment. Finally, the subject’s skin must be monitored during equilibration with a suitable IRT to measure the equilibration achieved.

### 2.4 Selection of Reference Body Measurement Site and Febrile Threshold

Medical thermometry devices have some latitude in specifying a febrile threshold for their device outputs. Some NCITs and IRTs specify thresholds as low as 99.5F. This work is concerned with assessing clinical suitability for febrile detection and because there are several body sites and thresholds to choose from a choice must be made for the reference site and febrile threshold. We aimed to select a site that balances accessibility and clinical accuracy and furthermore to select the most commonly used febrile threshold for this site and thus, we selected oral thermometry and 100.4F as the febrile threshold in this work. A deeper discussion of device-dependent thresholds is outside the scope of this study, but there are serious potential negative repercussions from relying on a lower threshold to ameliorate the loss of sensitivity from a strong bias-to-normal algorithm. Realistically, users will often ignore a reading between 99.5 and 100F or even higher.

It is important to point out the general concept of elevated body temperature detection has additional complexities we will not delve into deeply but briefly touch on a few here. Normal physiologic variability in core temperature is well established, with the time-of-day and menstrual cycle generally accepted as contributing the largest sources of variability, each ranging from 0.3 to 1F in magnitude{31}. The idea of an adjusted “average” body temperature, lower than the value of 37C or 98.6F arrived at in the 19th century, is still of considerable debate. Several possible causes or confounds are being investigated, such as increases in medication usage, changes in diet and lifestyle or decreases in latent infectious disease. However, accurate body thermometry is not simple, there is a range of rigor to these studies such as mining clinical records that may have been recorded with a variety of methods such as NCITs, and the view of some clinicians with routine involvement in close monitoring of body temperature (anesthesiology) is that 37C (98.6F) is a representative average{25, 31}. Ultimately it is not at all clear whether there is such a change.

## 3 Methods

### 3.1 Subjects and Test Summary

Under supervision of a St Olaf College’s IRB, we recruited 28 individuals (mean age 22.25 ± 7.45 yrs, 159.7 ± 36.5 lbs, 5.54 ± 0.32 ft, 12 males (43%); males mean age 23.58 ± 10.76 yrs, 161.6 ± 33.4 lbs, 5.75 ± 0.31 ft; females mean age 21.25 ± 2.84 yrs, 158.3 ± 36.5 lbs, 5.38 ± 0.22 ft) at St Olaf College from the student and faculty populations and, in five 2-hour sessions staggered over 8 days in August 2021, obtained oral temperature measurements and elevated temperature measurements after each individual stood for 10 minutes in each of four temperature-controlled tents, while being scanned with a thermal camera. After the 10 minutes concluded, the individual would stand at the rear wall inside the tent, behind a removable insulated cover and radiative shield, and through which they could be oriented towards a rotating table containing several secondary thermographic systems and the Device #3 thermometer. The tents were arranged around the rotating table (described below and in Fig. 9), configured to achieve the same distance and orientation of each system to each subject within an inch (tape measure to center console) for each tent, with markings indicated on the floor for rotational positioning. Each tent contained a space heater and relay controller configured for a 1F window about the following air temperatures: 76F, 80F, 84F, 88F (setpoints were approximate, each tent was monitored continuously via its own IRT device taking canthi measurements and each having an air temperature sensor calibrated to an accuracy of 0.1F).

**Fig. 9.**
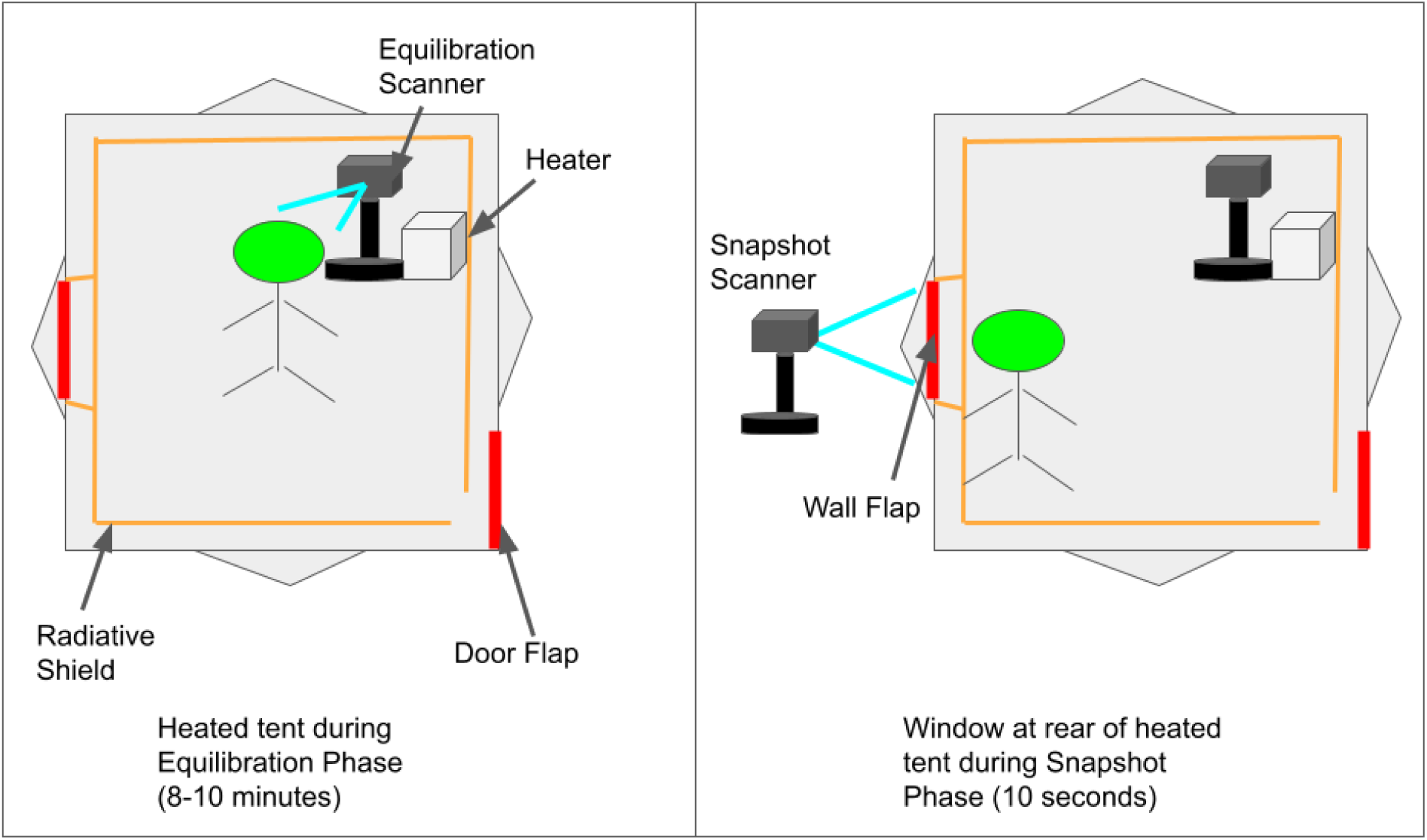
Environment (tent) with control of air temperature using a relay-controlled space heater. Equilibration is monitored using an *equilibration* scanner (IRT) acquiring sequential images over 10 minutes. NFC reader embedded in wall to activate equilibration scanner prior to entry of chamber and start of equilibration to synchronize acquisition of images and air temperature. Tent is modified for a second unoccluded pathway into the environment via a wall flap that is opened for a second NFC read and scan with scanner under test (“*snapshot*” scanner) and testing with other secondary scanners.

**Fig. 10.**
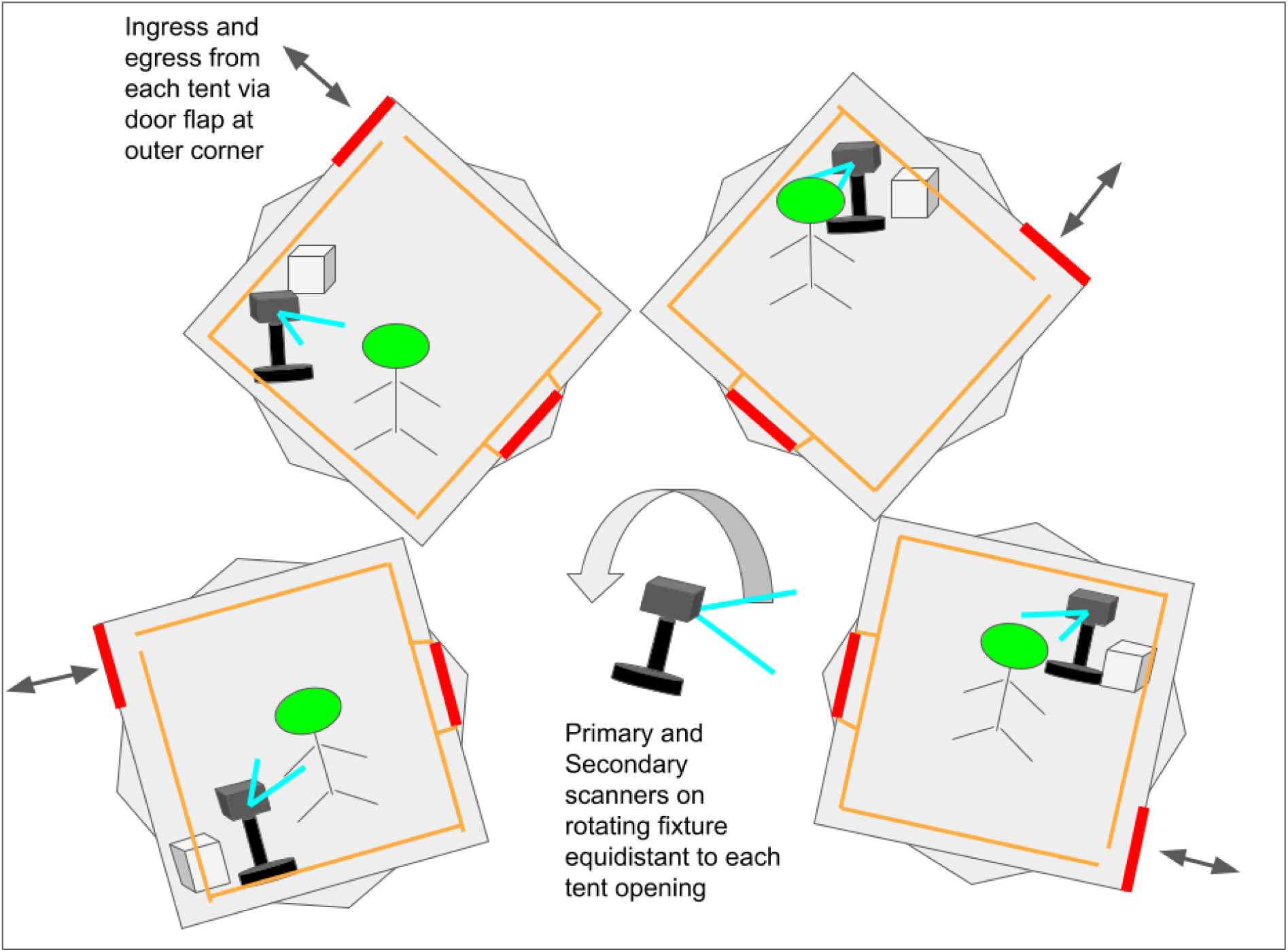
Elevated temperature environments arranged uniformly about the scanners sited on a rotating lectern. Each environment is set to a different target temperature. A 4-foot tall lectern on freely rotating casters (the snapshot acquisition fixture) was used to hold the devices under test, with tape placed on the floor indicating the outlines of the lectern in order to maintain a consistent distance and orientation with respect to each tent. Each tent was arranged about this lectern with the opening of each tent facing the lectern and perpendicular to this axis as shown in Fig. 10, such that the distance from a subject’s face within each tent opening was maintained at 5 feet from Devices 2 and 4 mounted on the lectern. Device 5 was mounted on a separate tripod to the left and behind the lectern, Device 1 was mounted on a separate tripod to the right side of the lectern as shown in Fig. 8.

**Fig. 11.**
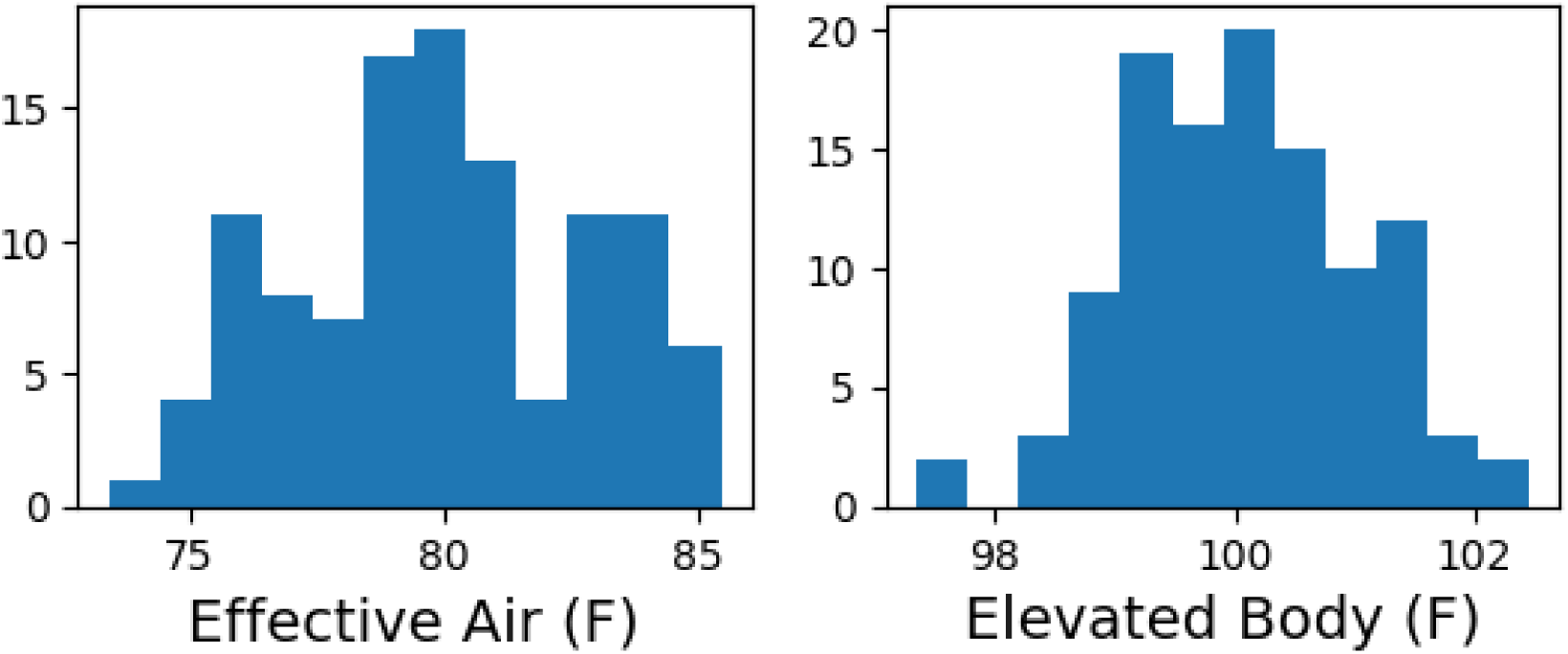
Tent effective air temperature and simulated elevated body temperatures.

**Fig. 12.**
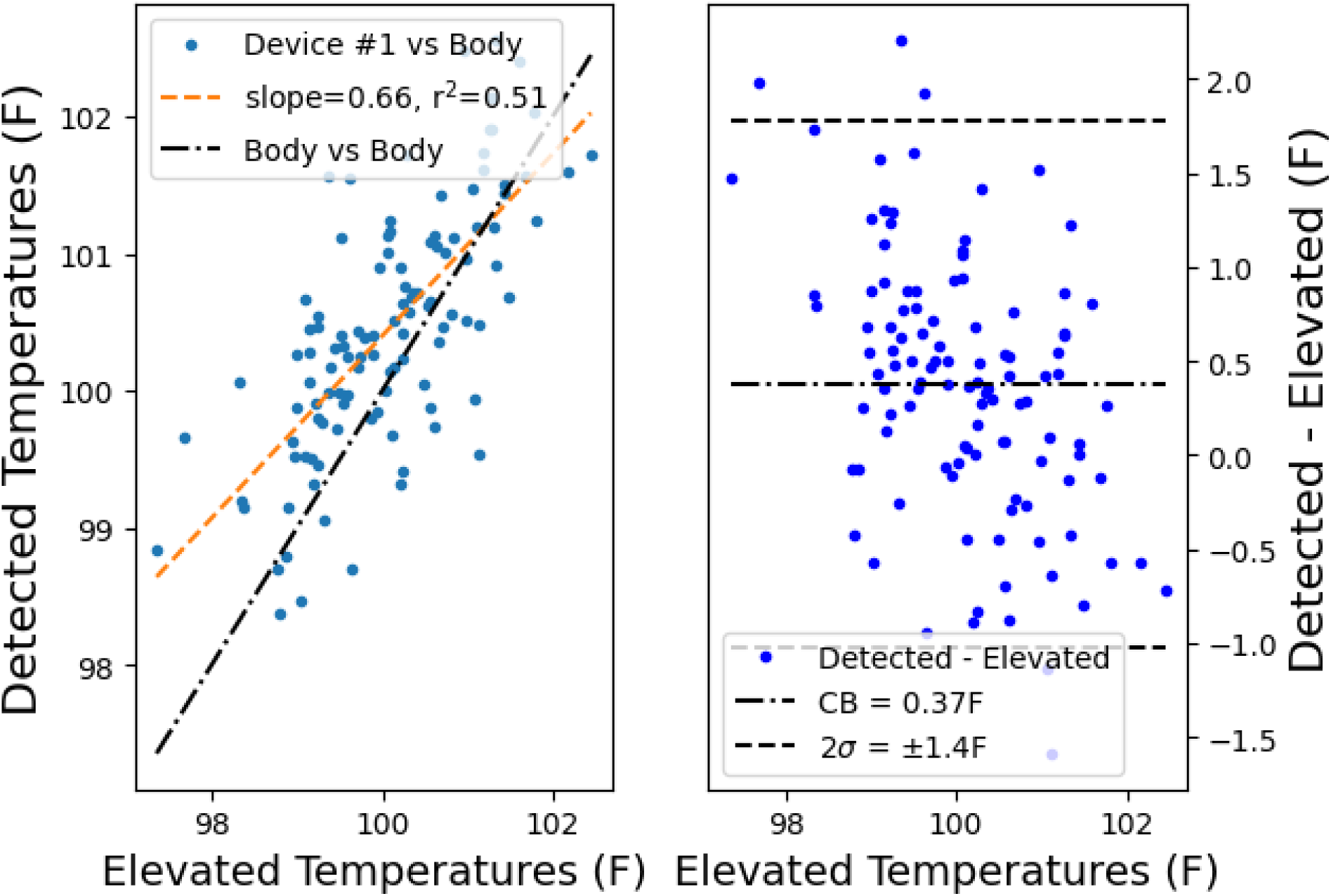
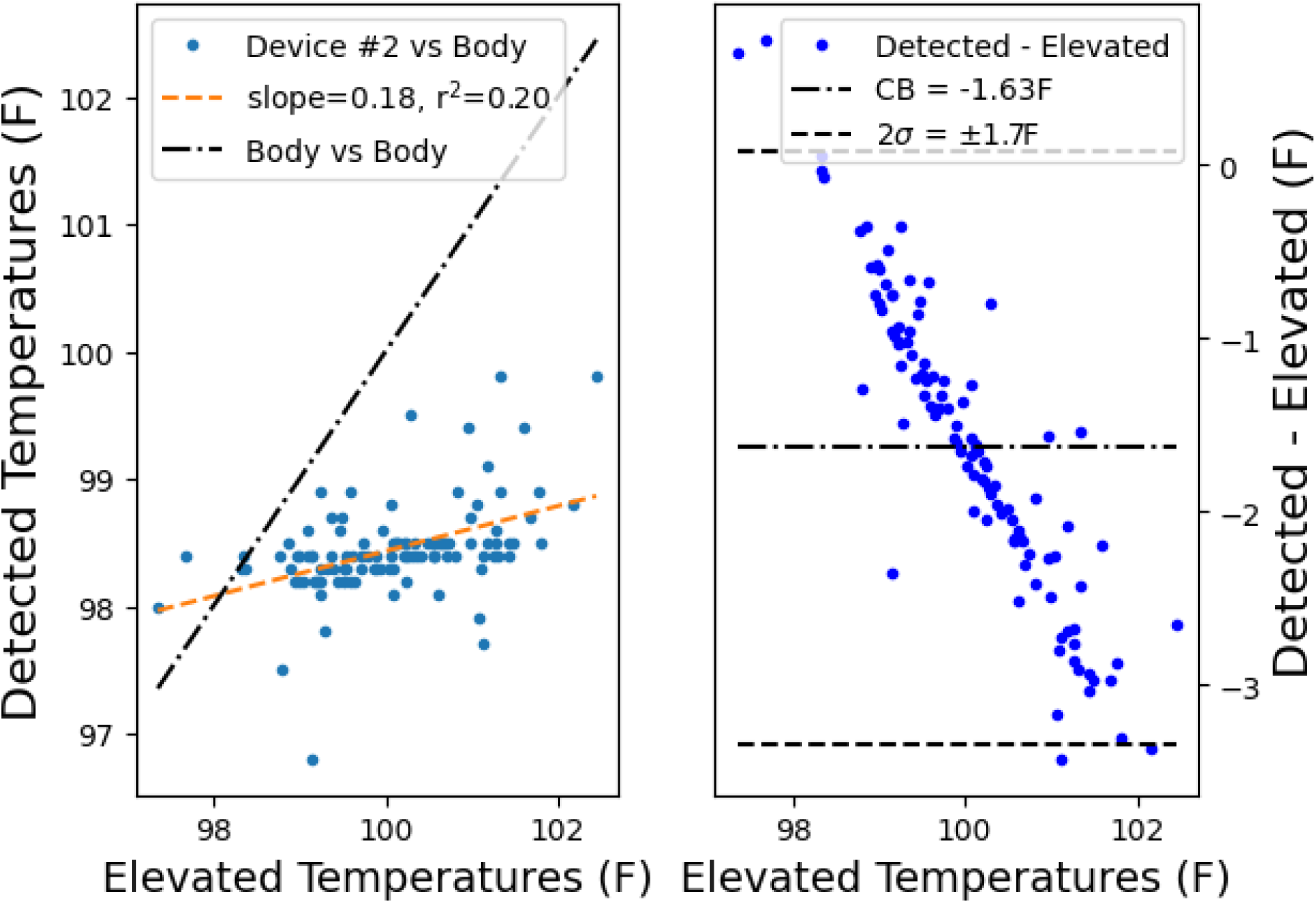
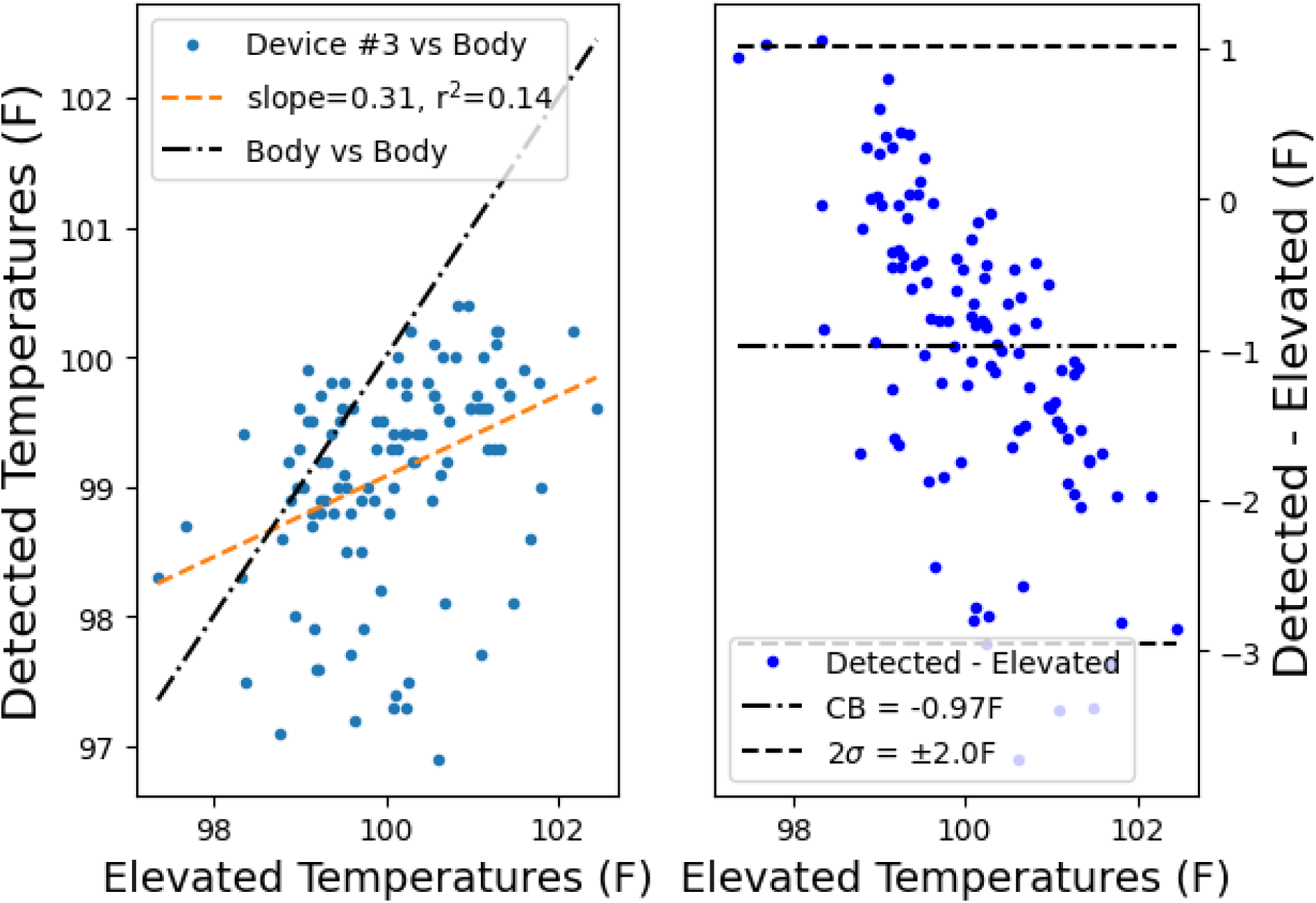
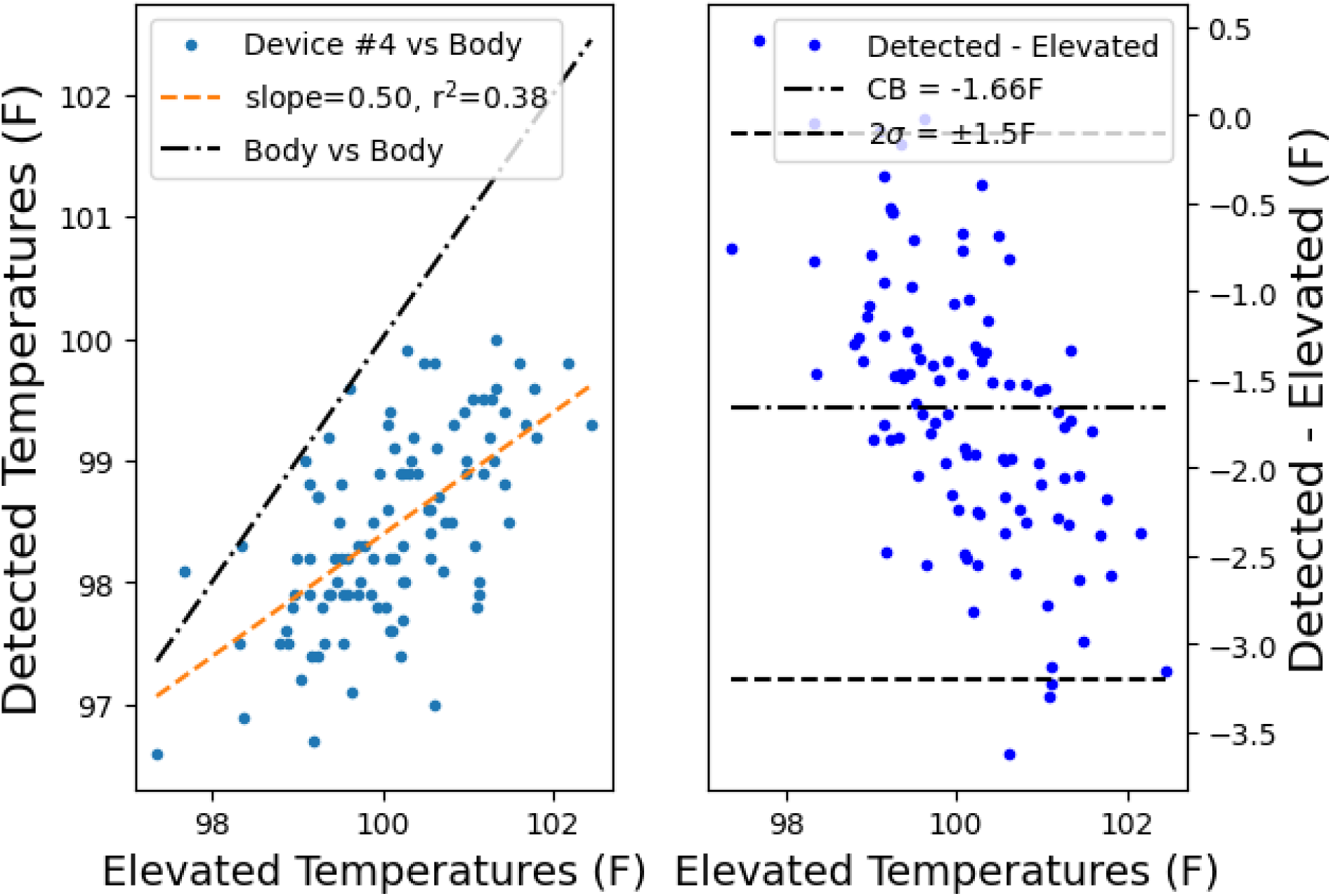
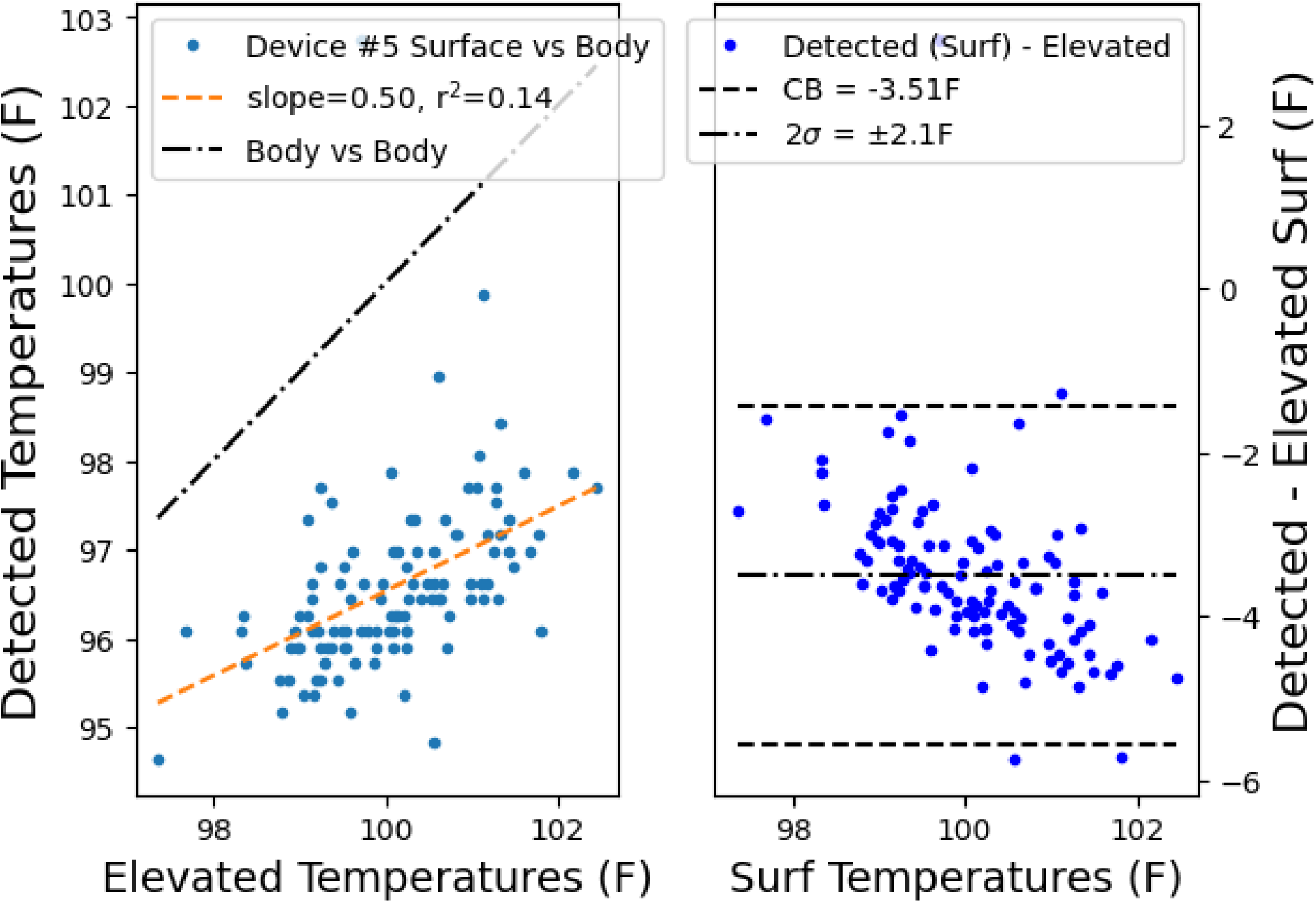
Left column shows detected vs simulated elevated temperatures and right column shows Bland-Altman plot (using elevated temperatures instead of the average in the x-axis due to potential bias in the detected temperatures), for each of Devices #1-5.

### 3.2 Thermometry Devices

Oral thermometry was performed using a clinical heated probe oral thermometer (Welch Allyn SureTemp Plus 690) which had its accuracy tested in an immersion circulator water bath accurate to 0.1C traceable to NIST calibration standards (PolyScience SD7LR-20). This oral thermometer has a quick-read mode, returning an estimated core temperature in under 10 seconds, however this mode biases towards the population average (See Supplement Fig. S3), so for clinical study this thermometer must be switched into a direct mode and the operator must wait for several time constants, which in this study was taken to be 2 minutes past the quick-read result (approximately 2.25 minutes). Subjects and operators were trained on proper preparation and placement of the oral probe to the sublingual pocket using the manufacturer’s printed operating instructions.

The non-contact devices included four IRTs and one NCIT, as shown in Table 2, with each device’s specified (claimed) accuracy, febrile alarm threshold and approximate time required to obtain a temperature. Note, Device #5 operates only in surface temperature mode, and furthermore relies on a +1C (1.8F) threshold, based on the average of the last up-to-8 subjects within a time window. Because the moving average of subjects would be biased upwards by the elevated temperatures seen, the moving threshold was not used. Testing of the device indicated an average of 35.1C with non-elevated face temperatures in ambient temperatures of 22C, thus a threshold of 36.1C was used for Device #5 in subsequent analyses.

The Device #1 devices inside the tents and at the center testing station were configured to start measurement once the first face was detected after a USB-attached near-field communication (NFC) reader was activated by a valid NFC sticker. Each face measurement was initiated at a regular interval (minimum of 3 seconds after the previous for snapshot measurements at the end of the 10 minute equilibration and minimum of 4 seconds for equilibration measurements, and in each case requiring a further second to complete the face measurement), and relied on the product face measurement algorithm which records unix timestamps, distance to target, ambient temperature, inner canthus surface temperature, calculated body temperature, and various system statistics was modified to save additional data not normally recorded for every face measurement, including a secondary 0.1F-calibrated ambient temperature probe, face detection bounding boxes, and the set of thermal images used in each measurement. All Device #1 devices were synchronized via NTP and were monitored from a laptop at the center console which also operated the Device #4 Scan software (version 1.9.1.0).

### 3.3 Testing and Adjusted Environments

The testing environment was a 3rd floor college lecture room with concrete flooring and insulative walls and ceiling, with four environmental chambers arranged around a testing station. The testing room radiative temperature was approximately within 1C of the ambient air temperature, which was maintained at 71.2±0.73F (during each test session the air temperatures were within 0.56 to 1.46F of a setpoint). The environmental chambers consisted of a 300 denier nylon free-standing tent with no floor (THUNDERBAY Ice Cube 3 Man Portable Ice Shelter), one zippered entrance flap at a corner and four removable windows, three of which were covered and one of which was modified to accommodate a cover designed for easy removal.

Each tent contained a portable space heater (Andily Electric Space Heater at 750W setting) controlled by an on/off relay system (Ketotek KT4000) configured to operate with a 1F window below the setpoint. Setpoints were arranged to approximate a range of elevated body temperatures from 99.5F to 102.5F (see github repository for additional photographs of the test environments). The temperature probe was suspended approximately above where the subjects head would be positioned (between 6” and 1 foot of the ceiling), intended to avoid gradients due to the tent itself or to any vertical air column gradients within the tent. Radiative shielding (Mybecca 100% unbleached Cotton Muslin Fabric) hung from the ceiling edges plus insulation above the ceiling and on the concrete floor are used to more closely match the effective (radiative background) temperature to the actual air temperature inside the tent. A ¼” polycarbonate trapezoidal frame was adhered to the tent over the wall flap opening and a ¼” foam-core removable cover was designed to insert into this opening. The radiative shield over the wall flap was cut to match the bottom 3 sides of the trapezoidal wall flap opening and the bottom of the radiative shield pinned to the tent at the bottom of the wall flap opening so that volunteers could access the window.

### 3.4 Study Procedures

Each subject was given a data collection sheet having an NFC sticker and alphanumeric indicator (incrementing the letter by study group and incrementing the number by individual, e.g. C1 through C8 refers to the 8 subjects being acquired during the third study group). This sheet was used to collect demographic and potential confound information and the entries for oral thermometry acquired before the start of the study and oral thermometry in quick-read and monitor mode after each equilibration session. After completing their demographic information, each subject practiced using a dedicated training scanner having a live view of the thermal images. After practice was complete, the first subject (e.g. C1) was given a tent to proceed to (e.g. “volunteer C1 please proceed to tent A”), at which point the subject scanned the NFC sticker on their sheet on an NFC reader embedded in the outer wall prior to entry (note, the first five groups were acquired without a practice session and with the NFC reader inside the tent, which led to uncertainty between the start of equilibration and the first image acquired during equilibration). After entering the tent, assisted by study personnel, the subject stood in front of the equilibration scanner for 10 minutes until its display indicated completion, which was also monitored remotely by personnel at the snapshot acquisition fixture. On completion, personnel prepared by orienting the fixture to the completed tent, placed the reference blackbody for Device #4 on the wall frame, before removing the wall frame cover and passing the second NFC reader under a cutout in the radiative shield inside the tent. The subject scanned their data collection NFC sticker on this NFC reader and the personnel lifted the radiative shield cutout, instructed the subject to stand just inside the opening, and personnel then proceeded to acquire a single acquisition using each scanner being tested. Device #1 automatically acquires data once a face is observed and the snapshot scanner was configured to acquire three snapshots. Devices #2 and #5 both continuously report a body temperature and surface temperature respectively. Device #4 automatically reports a body temperature once a face is observed with the blackbody in view next to the face. Device #3 (NCIT) was manually operated by study personnel by swiping the probe head across the subject’s forehead-to-temple region following manufacturer’s instructions and sanitized with an EPA-approved (List N for COVID) disinfecting wipe after use. All temperatures except for the Device #1 were recorded manually by study personnel. Once the test snapshots are complete, the subject is directed to exit the tent via the entrance flap and receive an oral thermometry measurement, with both the quick-read and a two-minute monitor-mode temperature recorded. After all four tent acquisitions and oral temperatures, subject participation was complete.

### 3.5 Study Analyses

For each equilibration dataset, the inner canthus, background radiance and calibrated ambient air temperatures were filtered with a 3-wide, 1st-order Savitsky-Golay filter over time and re-aligned to a unified end-of-equilibration time grid using the unix timestamps for each measurement. Background radiance was estimated using the median of the coldest 10% of pixels, verified in each measurement to correspond to the radiative shield material. Inspection of the ambient temperatures revealed a difference between the air temperatures and background radiance, the effect of which was approximated using a weighted sum of radiative and ambient air temperatures in order to derive an effective environmental temperature timecourse (see Supplement - Radiative and Effective Temperature).

Simulated elevated body temperatures were calculated using the inside-tent measured inner canthi surface temperature at the conclusion of equilibration, prior to exposure to the outside-tent scanning devices. In this method, the equilibrated surface temperature is inserted into Equation 1 with the ambient outside-tent air temperature to obtain an ***simulated elevated body temperature***.

For each Device #1 snapshot dataset taken from the outer testing environment, the three inner canthus measurements were taken over 8 seconds (every 4 seconds) and the average was used to calculate the Device #1 ***detected body temperature*** using the outer testing environment’s air temperature. For the Device #5 scanner, the reported ***detected facial surface temperature*** was recorded without any extrapolation. All other devices reported a ***detected body temperature***.

For each device’s manufacturer-specified febrile threshold (100.4F for Devices #1 and #4, 99.14F for Device #2, 100.1F for Device #3, and an approximation of the moving-average threshold for Device #5), the true positive rate (TPR) was determined from the percentage of simulated elevated body temperatures over the device’s threshold (number of febrile) that were detected as above the device’s threshold (detected as febrile). The false positive rate (FPR) was determined by dividing the number of below-threshold simulated body temperatures that were detected above the device’s threshold (false positives) by the total number of below-threshold simulated body temperatures (number of non-febrile). The detected body temperature output versus the simulated elevated body temperature was plotted for each device (using surface instead of body temperatures for Device #5 only), and a linear fit performed, with the fit coefficients inspected for under- or over-estimation of body temperatures. Bland-Altman plots of detected minus simulated elevated temperatures versus simulated elevated temperatures were plotted for each device with 2 sigma bounds, or limits of acceptability (LA) and the average difference, or clinical bias (CB), denoted by dashed lines.

Note, Device #5 calculates the moving average of the last 8 subjects and sets a changing threshold of 1C greater than this average. This threshold would be manipulated by the presence of elevated face temperatures, whether simulated or actual, and thus cannot be used to assess efficacy without a pool of subjects with normal body temperatures on standby to continually reset the moving average, which was not possible with the limit on the number of subjects allowed in the testing environment in the present study. To compare devices we calculated the surface temperature corresponding to the average oral temperature of the study population and added 1C (1.8F) to use as the selection threshold to identify simulated febrile or non-febrile, and to obtain the device detection threshold we used Eq 2 to extrapolate a surface temperature for the average oral temperature and added 1C (1.8F). For all other devices, we used each manufacturer-specified febrile threshold as given in Table 2.

## 4 Results

### 4.1 Oral and simulated elevated temperatures

The oral temperatures were averaged post-tent exposure (the average oral temperatures per tent, in increasing tent temperature order, were 98.33F, 98.31F, 98.33F and 98.45F) and no trend was observed in oral temperatures after elevated air exposure. Note however the average oral thermometry in the last exposure was 0.12F greater (2.28 standard deviations) than the average of the oral thermometry after the first three exposures.

Twenty-eight subjects in four chambers produced 112 elevated body temperatures (one measurement corrupted), enumerated in Table 3 and in Supplemental Table S1.

**Table 3.** Subject mean and standard deviation (across four measurements) oral and simulated elevated body temperatures at the conclusion of equilibration for each tent, with elevated body temperatures based on the surface temperature measured inside the tent, Eqn 1 and the testing room ambient temperature.

| Subject | Oral - Quick | Oral - Long | Tent A | Tent B | Tent C | Tent D |
| --- | --- | --- | --- | --- | --- | --- |
| E1 | 98.15±0.15 | 98.28±0.15 | 98.80 | 99.28 | 99.00 | 100.57 |
| E2 | 98.47±0.83 | 98.18±0.13 | 100.09 | 101.13 | 101.07 | 101.81 |
| E3 | 99.08±0.30 | 98.95±0.11 | 98.78 | 99.18 | 100.25 | 100.62 |
| E4 | 98.88±0.33 | 98.90±0.12 | 99.22 | 100.12 | 100.09 | 101.10 |
| F1 | 98.35±0.05 | 97.98±0.04 | 99.59 | 99.39 | 99.70 | 101.18 |
| F2 | 98.85±0.27 | 98.38±0.19 | 100.23 | 99.36 | 100.08 | 101.77 |
| F3 | 98.65±0.18 | 98.25±0.15 | 100.03 | 100.54 | 101.26 | 101.27 |
| F4 | 99.00±0.37 | 99.10±0.33 | 99.95 | 99.89 | 100.74 | 100.67 |
| F5 | 98.00±0.07 | 97.40±0.16 | 99.87 | 99.43 | 99.90 | 100.99 |
| F6 | 98.43±0.25 | 98.48±0.08 | 100.70 | 100.34 | 101.48 | 101.68 |
| F7 | 99.20±0.21 | 98.93±0.19 | 100.41 | 101.44 | 101.05 | 102.17 |
| G1 | 98.28±0.13 | 98.10±0.25 | 99.25 | 99.72 | 99.97 | 100.63 |
| G2 | 98.90±0.07 | 98.88±0.04 | 98.36 | 98.95 | 99.58 | 99.75 |
| G3 | 98.23±0.15 | 97.65±0.17 | 98.86 | 99.32 | 100.49 | 100.57 |
| G4 | 99.18±0.29 | 98.50±0.16 | 97.68 | 98.34 | 99.15 | 99.09 |
| G5 | 98.08±0.13 | 97.55±0.18 | 98.33 | 99.14 | 99.15 | 99.51 |
| G6 | 98.58±0.11 | 97.95±0.18 | 99.04 | 99.46 | 99.09 | 100.65 |
| H1 | 98.15±0.23 | 98.35±0.05 | 99.36 | 100.13 | 101.33 | 100.97 |
| H2 | 98.20±0.21 | 98.10±0.08 | 99.55 | NA | 101.43 | 100.61 |
| H3 | 98.30±0.12 | 98.55±0.15 | 100.07 | 100.21 | 101.27 | 100.61 |
| H4 | 99.70±0.61 | 99.50±0.12 | 101.19 | 101.59 | 102.45 | 101.34 |
| I1 | 98.38±0.15 | 97.75±0.05 | 100.21 | 100.25 | 100.56 | 101.12 |
| I2 | 98.60±0.14 | 98.43±0.04 | 99.00 | 99.25 | 100.81 | 100.24 |
| I3 | 98.68±0.08 | 98.75±0.05 | 99.23 | 99.62 | 100.36 | 100.30 |
| J1 | 98.33±0.29 | 98.05±0.15 | 99.53 | 99.48 | 100.83 | 100.07 |
| J2 | 98.95±0.29 | 98.98±0.11 | 99.16 | 99.64 | 99.54 | 100.27 |
| J3 | 98.68±0.49 | 98.45±0.15 | 99.80 | 100.30 | 101.32 | 100.15 |
| J4 | 98.10±0.19 | 97.55±0.09 | 97.35 | 98.89 | 99.24 | 98.98 |
| **All** | **98.58±0.41** | **98.35±0.51** | **99.42±0.8** | **99.61±0.8** | **100.40±0.9** | **100.68±0.8** |

### 4.2 Detected versus simulated elevated temperatures

These figures show what may be bias-to-normal behavior in devices 2 and 3. The ideal Bland-Altman plot should show no trendline, but in these cases, a negative slope in the Bland-Altman plot for devices #2 and #3 indicates that as the object temperature increases, the reported temperature will tend to the same value. Note, we did not plot the difference versus the average because in the presence of a bias-to-normal, the average could obscure these effects.

The results shown in Table 4 are similar to those produced in a recent study{48} that used oral thermometry, IRTs and careful adherence to ISO guidelines. In particular, Figure 3 of that study shows the IRT vs oral temperatures for three IRT systems, each of which show a narrower range of IRT-reported body temperatures compared to the oral temperatures. Of note, Camera 2 is the same device and model as Device #5 in the present study, and its surface temperature output found to fit the body temperatures with a similar squared correlation coefficient and slope with a slope of 0.5 here versus 0.6186 in {48}, both of which refer to linear fitting of the surface temperature output, not the calculated body temperature. Of note, the present study did not use a felt board behind the subject for Device #5 due to the tent design, which may have contributed to this difference (see figure 1 in {48}). The average temperatures detected by each device is shown in Table 5, with the average elevated temperature for each tent for comparison.

**Table 4.** Linear fit slope coefficient for each device’s detected body temperature (or converted via Eqn 1, for Device #1 and #5) versus elevated body temperatures, r-squared correlation between detected and elevated, TPR, FPR, CB and LA. Device #5’s CB was not assessed.

|  | Slope | r<sup>2</sup> | TPR | FPR | CB | LA |
| --- | --- | --- | --- | --- | --- | --- |
| Device 1 | 0.66 | 0.51 | 0.85 | 0.34 | 0.37 | 1.4 |
| Device 2 | 0.18 | 0.20 | 0.05 | 0 | -1.63 | 1.7 |
| Device 3 | 0.31 | 0.14 | 0.11 | 0 | -0.97 | 2.0 |
| Device 4 | 0.50 | 0.38 | 0.0 | 0 | -1.66 | 1.5 |
| Device 5 | 0.50 | 0.14 | 0.82 | 0.28 | NA | 2.1 |

**Table 5.** Average simulated elevated temperatures per tent, average detected body temperatures by device per tent for Devices 1-4, average detected surface temperature for Device 5.

|  | Tent A | Tent B | Tent C | Tent D |
| --- | --- | --- | --- | --- |
| **Body (F)** | **99.15** | **99.61** | **99.79** | **100.45** |
| Device 1 | 99.9 | 100.15 | 100.68 | 101.03 |
| Device 2 | 98.3 | 98.41 | 98.45 | 98.62 |
| Device 3 | 98.84 | 99.05 | 99.23 | 99.29 |
| Device 4 | 97.93 | 98.26 | 98.69 | 98.87 |
| Device 5 | 95.93 | 96.47 | 96.93 | 96.94 |

Note the distribution of elevated temperatures straddled the febrile thresholds used and as a consequence, the false positive rate can be expected to be high for the thresholds used here, yet the real-world false positive rate would be expected to be far lower when it is weighted by the distribution of real temperatures near the threshold.

## 5 Discussion

The intent of this study was to bring attention to the methods used for non-contact body thermometry and reveal serious shortcomings in what is widely used today as a public health screening tool. This study revealed systematic differences in performance between devices by demonstrating and using a new test method to measure real-world sensitivity. This test relies on objectively manipulating the surface temperatures, and therefore the calculated body temperatures reasonably expected, based on established findings in physiology and patent literature and the underlying method for febrile detection. The linkage between elevated skin and elevated core is central to all of non-contact febrile screening and thus it is difficult to object to this method wholesale without also objecting to all of non-contact thermometry. NCITs and IRTs are widely used for febrile screening and obtaining vital sign assessments, and the requirement that such devices be tested on febrile patients limits innovation by manufacturers and independent assessment by users and regulators. Worryingly, the findings in this study echo recent discoveries of undisclosed “bias-to-normal” algorithms in a range of IRTs recently released in the months following the COVID-19 pandemic{47} under FDA guidance and we extend those findings to a variety of FDA-cleared non-contact thermometry systems and IRTs that have been widely used for febrile screening. This new test method can help device manufacturers, independent test laboratories and regulators to each do their part to improve the quality of non-contact thermometry.

Several limitations were identified during the performance of this study. First, the design did not include a pre-tent temperature snapshot (e.g. taken while the subjects had equilibrated to the testing room outside the tent environment), which would be an important addition and was an oversight. Secondly, the tent air temperature control achievable with off-the-shelf relay controllers was not sufficiently stable and the output vent of the space heaters was not attenuated to prevent noticeable convective flow within the tent, introducing variability into the equilibration curves as the heaters switched on and off over a 1 to 5 minute timescale and bifurcated the heat transport coefficient for body heat flowing through facial skin into heat transport to still air and heat transport to convective air. This was the largest technical limitation of the study and was identified as the source of several deviations from expected values, such as inverted radiative and air temperatures in two of the 112 datasets in Table S3. The code listing includes a design of modified air temperature control that overcomes both these limitations, which was regrettably not available when the present study data was collected. Thirdly, the time between completion of equilibration and snapshot acquisition was not carefully controlled, causing potential variability in the final temperatures reached at the time of snapshot acquisition. The 3 snapshots acquired over 8 seconds by the Device #1 showed only 0.03F decline on average, with an average standard deviation of 0.14F across the three snapshots, implying minimal equilibration to the test environment. However, one device, Device #4, had trouble detecting some faces, particularly in the first three sessions, leading to delays in several cases of up to 30 seconds, which was not assessed independently nor was this occurrence adequately recorded. Study personnel estimate that this occurred in approximately 20% of Device #4’s data. Extrapolating linearly from the Device #1’s measured decline of 0.03F, it is possible the surface temperature seen in these delayed Device #4 snapshots were decreased by over 0.11F, which translates to a calculated body temperature decline of 0.13F by the time the most-delayed Device #4 snapshots were recorded. Finally, the range of elevated temperatures was too narrow to determine a 90% sensitivity for most of the devices.

Several questions of interest were identified for follow-up study. The F-value of skin may vary from person-to-person, under various conditions and from site-to-site. In the present study, a distance correction was used to remove the effect of distance on surface temperatures of the inner canthus, and this distance correction may be different for different sites, so that data acquired in the present study was not ideal for evaluating different sites. In a follow-up study, we intend to acquire skin site measurements in different ambient conditions and at a small plurality of controlled distances. Additionally, a secondary IRT system with good control over SSE and smaller spot sizes, such as Device #5, would be useful in obtaining independent measurements.

Time and again, the availability of gold-standard data, data that can be independently verified or controlled, has led to important breakthroughs in areas stuck on subtle systematic biases present in nearly all data at hand until that point. Given systematic biases that have in some cases taken months or even years of development effort in order to minimize these below acceptable levels for highly-advanced groups such as those working on space-based LWIR telescopes, it is not unlikely more surprises await the field as we iterate to an acceptable body temperature accuracy. To this end, it could be very beneficial to direct some focus to the development of accurate, controllable tissue phantoms for the realistic simulation of face temperatures. Ideally this would also be designed to match the dynamic temporal evolution of face temperatures by providing what could effectively appear as controllable *time constants* with spatial variation, which can be manipulated to match a variety of measured time constants. This could be a useful target for funding agencies concerned with improving preventive medicine or pandemic preparedness.

We cannot conclude this study without a simple observation: while it may seem reasonable that one could re-obtain good sensitivity on all these devices by using far lower thresholds, it is not as simple as reducing the threshold because that reduced threshold may only be there to compensate for an aggressive bias-to-normal behavior. Most importantly, the ambient temperature dependence of Devices #2-5 was not probed in this study. Had we obtained all snapshot measurements inside a warmer environment, the correct result would be normal (or closer to normal), while these devices would be reporting their somewhat higher body estimates because they do not measure or correct for ambient temperature in their application of an offset adjustment. The test environment maintained a very narrow range of ambient temperatures during the test, and real-world conditions must be assumed to be more variable than these stringent requirements. Over the past year, nearly everyone has seen IRTs or NCITs being operated in far more variable conditions. If customers merely reduce their thresholds to what can be expected to have good sensitivity, as soon as the ambient conditions change by 4C, depending on the direction, either the true positive rate will fall dramatically for typical low fevers or the false positive rate will rise over 50%. The natural tendency of an operator faced with a sequence of false positives is to increase the threshold until these false positives disappear. This is simply untenable, and the lack of an ambient temperature control is clearly a massive oversight by the field, which must be rectified before more damage is done. Methods such as relying on a moving average of the last N subjects, such as the algorithm used by Device #5, is hugely problematic and creates new issues that make it far harder to understand the efficacy of the device. Does that method work, is a question of great interest, and a simple simulation reveals that this can result in some, but limited, improvement over not compensating for ambient temperature because it becomes functionally blind for the first several subjects after a gap between the ambient temperatures during the last half of the subjects used in the moving average and the present subject. Typical fever inspection installations, for example an office environment reception area with 150 unique individuals scanned per day, have several gaps of 2-4 hours during the day, and ambient shifts of several degrees can occur between those periods. In half of those periods, the temperature is increasing, so the next two to four subjects will be likely to scan high, resulting in up to 16 subjects out of 100 the system is blind to and in that scenario will result in bursts of false positives, again leading most operators to manage by increasing the threshold. This process of adjusting the threshold for an acceptable false positive rate must be managed by the manufacturer of the device in some way, whether by directly engaging with the user to ensure a meaningful threshold is set or automating this threshold adjustment and reporting the effect of the adjustment on the sensitivity of the device to the user. However, this can be accomplished better by simply correcting for ambient temperature, which has a side benefit of being very low-cost.

### Disclosures

Dr Beall is a founder and CEO of Thermal Diagnostics LLC, which makes and sells the Device #1 IRT used in this study and Dr Beall has a financial stake in the company. Dr Schmidt is an Engineering Fellow at Fluke Corporation, which manufactures IRTs for *industrial* applications; his involvement in this research is independent of his work at Fluke Corporation and the views expressed here do not necessarily represent the views of Fluke. Dr Hinnerichs’ research is independent of the Department of Defense and does not reflect the opinion of the United States Army. Dr Adolph declares and attests she has no competing interests to declare. Mr Askegaard and Mr Berkesch are students at St Olaf College and declare and attest they have no competing interests.

## Data Availability

All anonymized data produced in the present work are available upon request to the authors and these data and the study materials (blank informed consent, data collection form and study protocol) is available in a public github repository at https://github.com/erikbeall/elevatedfacestudy2021.

## Acknowledgments

Study volunteers and testing locations for a pilot study in Summer 2020 and the use of Device #2 for the present study was provided by Cargill Incorporated. The testing room for the data collected in this study was provided by St Olaf College. Funding for an internship was provided by the Piper Center of St Olaf College. Other equipment and funding for volunteer remuneration was provided by Thermal Diagnostics LLC.

## Code, Data, and Materials Availability

All software, schematics and material lists are provided in a public GitHub repository at https://www.github.com/erikbeall/elevatedfacestudy2021.

## Author Biographies

**Erik Beall** received his PhD in Experimental Particle Physics from the University of Minnesota in 2005, and was Assistant Professor of Radiology in Cleveland Clinic Lerner College of Medicine until 2015, focusing on functional brain imaging methods development and application to neurodegenerative diseases and mood disorders. He is the author of more than a dozen journal papers and has written two book chapters. His current research interests include infrared calibration methods and modularization of thermal imaging sensors. He is currently the CEO of Thermal Diagnostics LLC and is a member of SPIE.

**Alden Adolph** received her PhD in Engineering Sciences from Dartmouth College in 2017. She is currently an Assistant Professor of Physics and the Director of Engineering Studies at St. Olaf College. Much of her research focuses on optical and thermal properties of snow, including the study of what factors lead to changing reflectivity of snow and assessing the accuracy of satellite-based remote sensing infrared surface temperature products in Greenland.

**Chris Hinnerichs** received his PhD in Epidemiology from Walden University, Minneapolis, Minnesota, in 2011, focusing on the efficacy of fixed infrared thermography for identification of subjects with Influenza-like illness. As a military medical service officer, his professional focus is Pacific disease surveillance, vector surveillance and mitigation efforts, applied research into the efficacy of mass spectroscopy proteomics for rapid identification of water pathogens, and past epidemiological investigations of the 2009 H1N1 pandemic, H5N1, MERS-CoV, mumps, and pertussis outbreaks.

**Matthew Schmidt** received his PhD in Condensed Matter Physics from the University of Minnesota in 1993. He is an Engineering Fellow at Fluke Corporation. He joined Fluke in 2005 when Fluke acquired Infrared Solutions, Inc. (“ISI”), Plymouth, MN. From 2005 thru 2010 he led Fluke’s Thermography Engineering team, with responsibility for developing Fluke’s handheld thermal imaging products and manufacturing capability. Since 2010 he has been responsible for innovation, technology and intellectual property development, primarily focused on Fluke’s thermal imaging business. Prior to Fluke’s acquisition of ISI, Dr. Schmidt was the VP of Engineering & CTO at ISI (2001-2005). His earlier roles include work as a Semiconductor Device Engineer at VTC Inc., and as a Senior Embedded Systems Engineer at Logic Product Development. He also co-founded and served as the President of MITI Corporation (later d/b/a OptiSat Medical, Inc.) from 1993 to 2000.

