## Supplement for "Principles and test methods of non-contact body thermometry"

**Appendix A: Supplemental Material**

*Calculating Simulated Elevated Temperatures from Oral Thermometry and Air Temperatures*

Equations 1 and 2 in Sec 2 can be combined to derive a simple relation for predicting the apparent elevated temperature resulting from equilibrating to one environmental temperature but being measured from a second, different environmental temperature, as

$$T_{core-simulated} = T_{core} + F * \left(T_{ambient\ equilibrated} - T_{ambient\ snapshot}\right), (3)$$

where $T_{core}$ is the core body temperature, $T_{core\text{-}simulated}$ is the apparent modulated temperature desired, $T_{ambient\ equilibrated}$ is the environmental temperature within which the subject has equilibrated for a sufficient time, and $T_{ambient\ snapshot}$ is the environmental temperature the device is being operated within. This relation can be used for determining useful elevated air temperatures for testing NCITs and IRTs. Eqn 3 can also be used for deriving the estimated elevated body temperatures (see below for this alternative method of obtaining simulated elevated body temperatures). Note in Sec 3 in the main body of this paper Eqn 1 and the measured elevated skin surface temperature was used for this purpose.

*Alternative elevated body temperatures*

An alternative means of estimating the elevated body temperatures that has less dependence on the use of Device #1 as a primary measurement system is to use Eqn 3 to obtain an ***alternative elevated body temperature***. It must be pointed out that the inside-tent canthi surface temperatures were obtained using a Device #1 device sharing the same calibration chain as the outside-tent Device #1 and therefore that data may be biased to perform better for the Device #1. However, the correlation coefficients were higher for all devices with the original method. This alternative method was examined here in the supplementary materials to provide an estimate less dependent on Device #1. These alternative elevated temperatures are shown below in Fig. S1 and Table S1. The temperatures obtained are broadly similar to those calculated with the other method yet there is less variability in the surface temperatures. Study personnel and subjects reported subjective feeling of moving air during heating phases, which is known to produce more-elevated surface temperatures than standing air. The heating air stations were located beneath the Device #1 equilibration scanner in each tent, which was the location of maximal convective flow. Thus, once the subjects moved to the rear wall and no longer experienced this level of convective flow, the surface temperatures may normalize somewhat but this was not measured and was a limitation to the tent design.

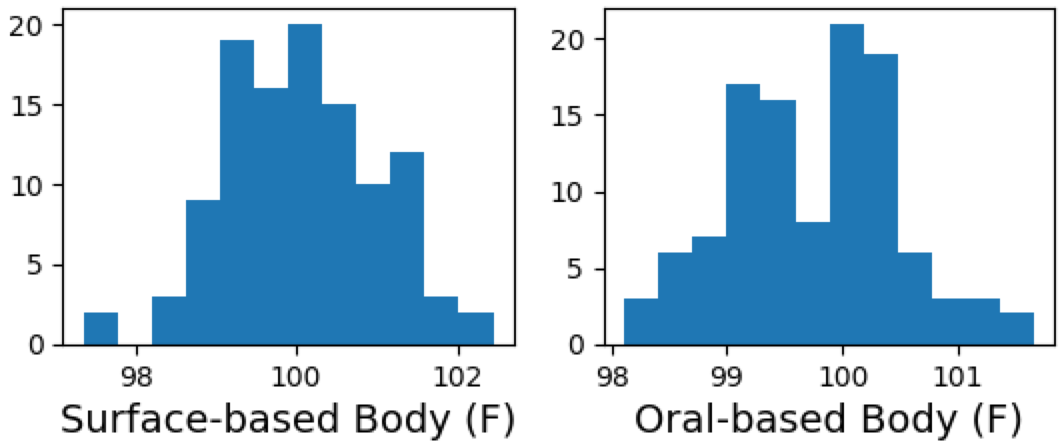


**Fig. S1** Calculated (simulated) elevated body temperatures by two methods. Surface temperature-based method used in Sec 4.1 shown to the left, oral thermometry-based method used here shown to the right.

**Table S1** Subject mean and standard deviation (across four measurements) oral and expected elevated body temperatures at the conclusion of equilibration for each tent, with elevated body temperatures based on the oral temperature, the ambient room air temperature and the tent effective temperature with Eq. 1.

| Subject | Oral - Quick | Oral - Long | Tent A | Tent B | Tent C | Tent D |
|---|---|---|---|---|---|---|
| E1 | 98.15±0.15 | 98.28±0.15 | 98.75 | 99.09 | 99.51 | 100.21 |
| E2 | 98.47±0.83 | 98.18±0.13 | 98.56 | 99.34 | 99.51 | 100.44 |
| E3 | 99.08±0.30 | 98.95±0.11 | 99.4 | 99.98 | 99.95 | 101.11 |
| E4 | 98.88±0.33 | 98.90±0.12 | 99.32 | 99.89 | 100.23 | 101.0 |
| F1 | 98.35±0.05 | 97.98±0.04 | 99.02 | 99.18 | 99.15 | 100.27 |
| F2 | 98.85±0.27 | 98.38±0.19 | 99.06 | 99.97 | 100.15 | 100.24 |

| | | | | | | |
|---|---|---|---|---|---|---|
| F3 | 98.65±0.18 | 98.25±0.15 | 98.76 | 99.81 | 99.92 | 100.29 |
| F4 | 99.00±0.37 | 99.10±0.33 | 100.19 | 100.1 | 100.49 | 101.55 |
| F5 | 98.00±0.07 | 97.40±0.16 | 98.55 | 98.55 | 98.73 | 99.22 |
| F6 | 98.43±0.25 | 98.48±0.08 | 99.16 | 100.02 | 100.08 | 100.73 |
| F7 | 99.20±0.21 | 98.93±0.19 | 99.83 | 100.01 | 100.43 | 101.31 |
| G1 | 98.28±0.13 | 98.10±0.25 | 99.12 | 99.1 | 99.01 | 100.44 |
| G2 | 98.90±0.07 | 98.88±0.04 | 99.46 | 100.33 | 100.22 | 100.65 |
| G3 | 98.23±0.15 | 97.65±0.17 | 98.11 | 98.68 | 99.22 | 99.96 |
| G4 | 99.18±0.29 | 98.50±0.16 | 99.07 | 99.58 | 99.88 | 100.33 |
| G5 | 98.08±0.13 | 97.55±0.18 | 98.34 | 98.87 | 98.59 | 99.36 |
| G6 | 98.58±0.11 | 97.95±0.18 | 98.28 | 99.37 | 99.12 | 99.9 |
| H1 | 98.15±0.23 | 98.35±0.05 | 99.12 | 99.65 | 99.9 | 100.44 |
| H2 | 98.20±0.21 | 98.10±0.08 | 98.79 | **NA** | 99.72 | 100.26 |
| H3 | 98.30±0.12 | 98.55±0.15 | 99.4 | 99.91 | 100.14 | 101.01 |
| H4 | 99.70±0.61 | 99.50±0.12 | 100.39 | 100.64 | 101.01 | 101.66 |
| I1 | 98.38±0.15 | 97.75±0.05 | 98.99 | 99.1 | 99.17 | 99.84 |
| I2 | 98.60±0.14 | 98.43±0.04 | 99.48 | 99.98 | 100.18 | 100.36 |
| I3 | 98.68±0.08 | 98.75±0.05 | 99.75 | 100.02 | 100.53 | 100.71 |
| J1 | 98.33±0.29 | 98.05±0.15 | 99.26 | 99.35 | 99.6 | 100.11 |
| J2 | 98.95±0.29 | 98.98±0.11 | 100.0 | 100.36 | 100.19 | 101.27 |
| J3 | 98.68±0.49 | 98.45±0.15 | 99.49 | 99.51 | 100.17 | 100.35 |
| J4 | 98.10±0.19 | 97.55±0.09 | 98.42 | 98.99 | 99.43 | 99.52 |
| **All** | **98.58±0.41** | **98.35±0.51** | **99.15±0.6** | **99.61±0.5** | **99.79±0.6** | **100.45±0.6** |

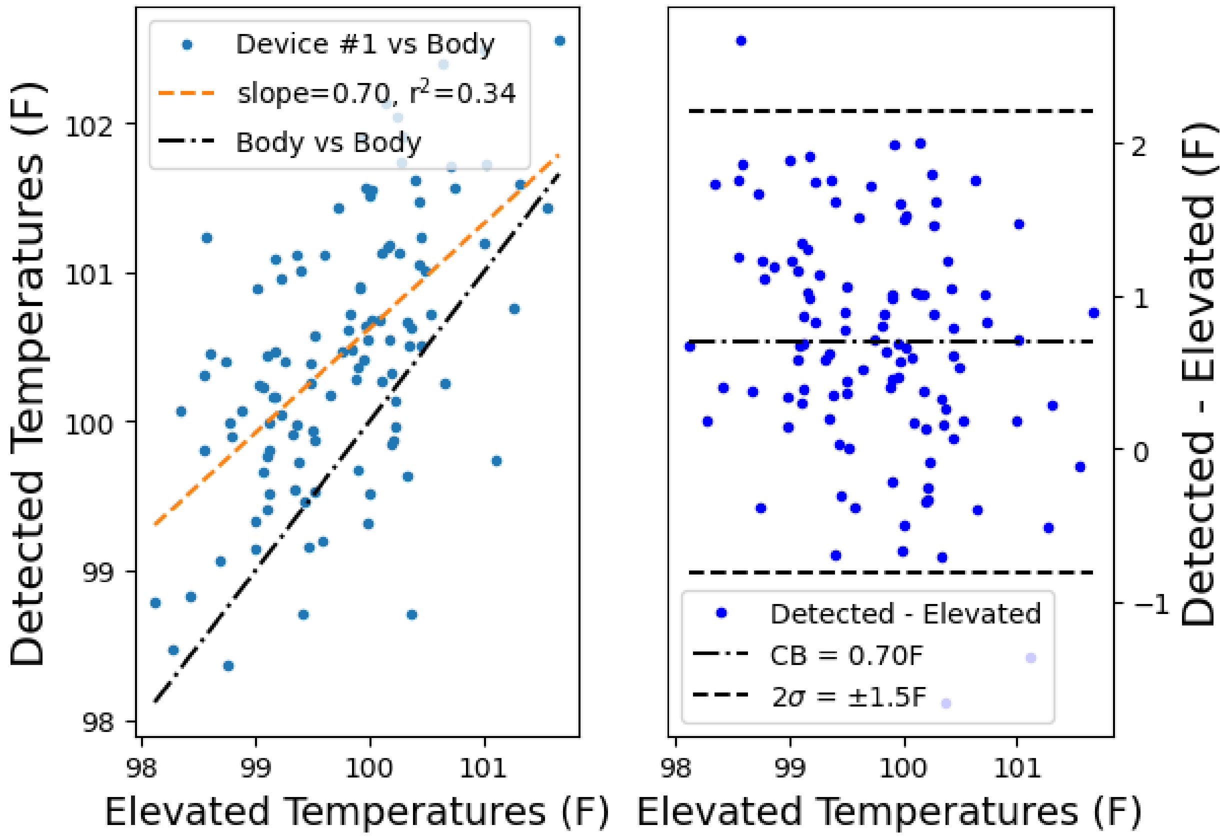
Device #1 vs Body
slope=0.70, r$^2$=0.34
Body vs Body
Detected Temperatures (F)
Elevated Temperatures (F)
Detected - Elevated
CB = 0.70F
2$\sigma$ = ±1.5F
Elevated Temperatures (F)
Detected - Elevated (F)

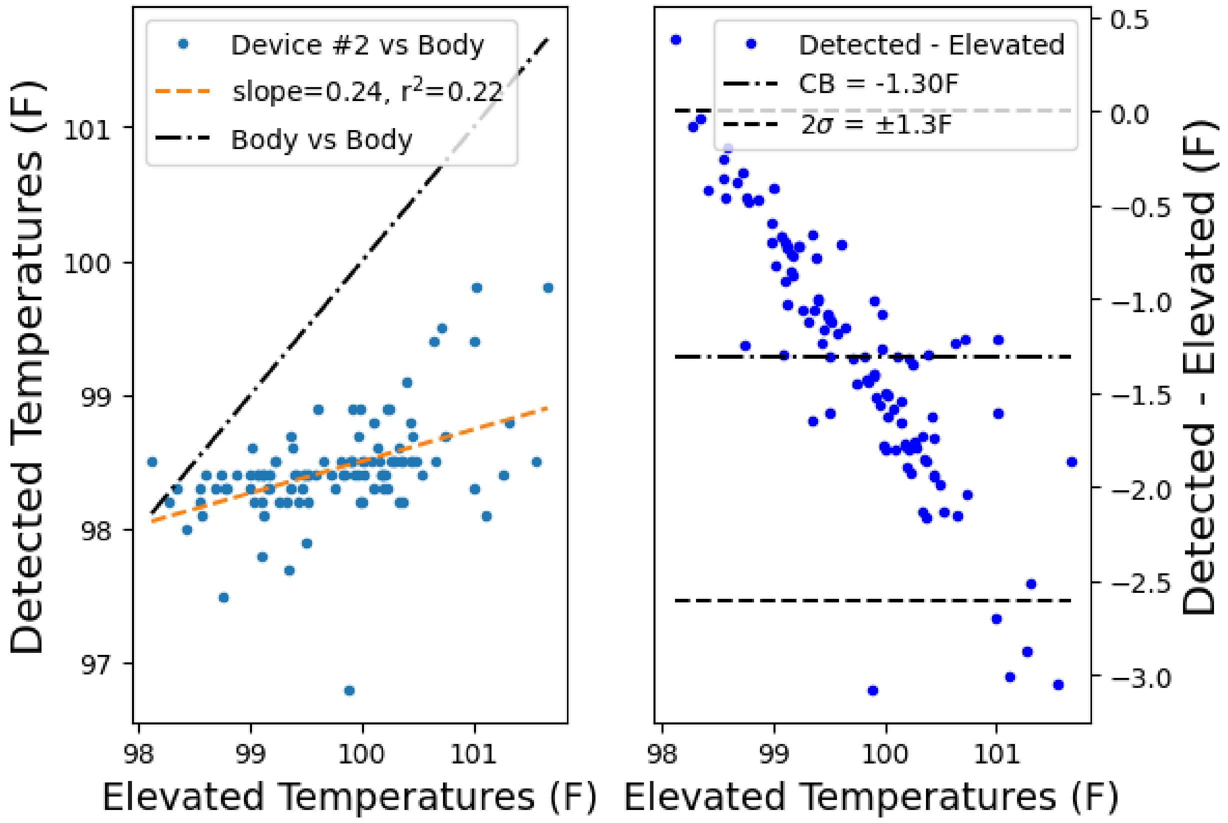

Device #2 vs Body
slope=0.24, r$^2$=0.22
Body vs Body
Detected Temperatures (F)
Elevated Temperatures (F)
101
100
99
98
97
98
99
100
101
Detected - Elevated
CB = -1.30F
2$\sigma$ = ±1.3F
Detected - Elevated (F)
Elevated Temperatures (F)
0.5
0.0
−0.5
−1.0
−1.5
−2.0
−2.5
−3.0

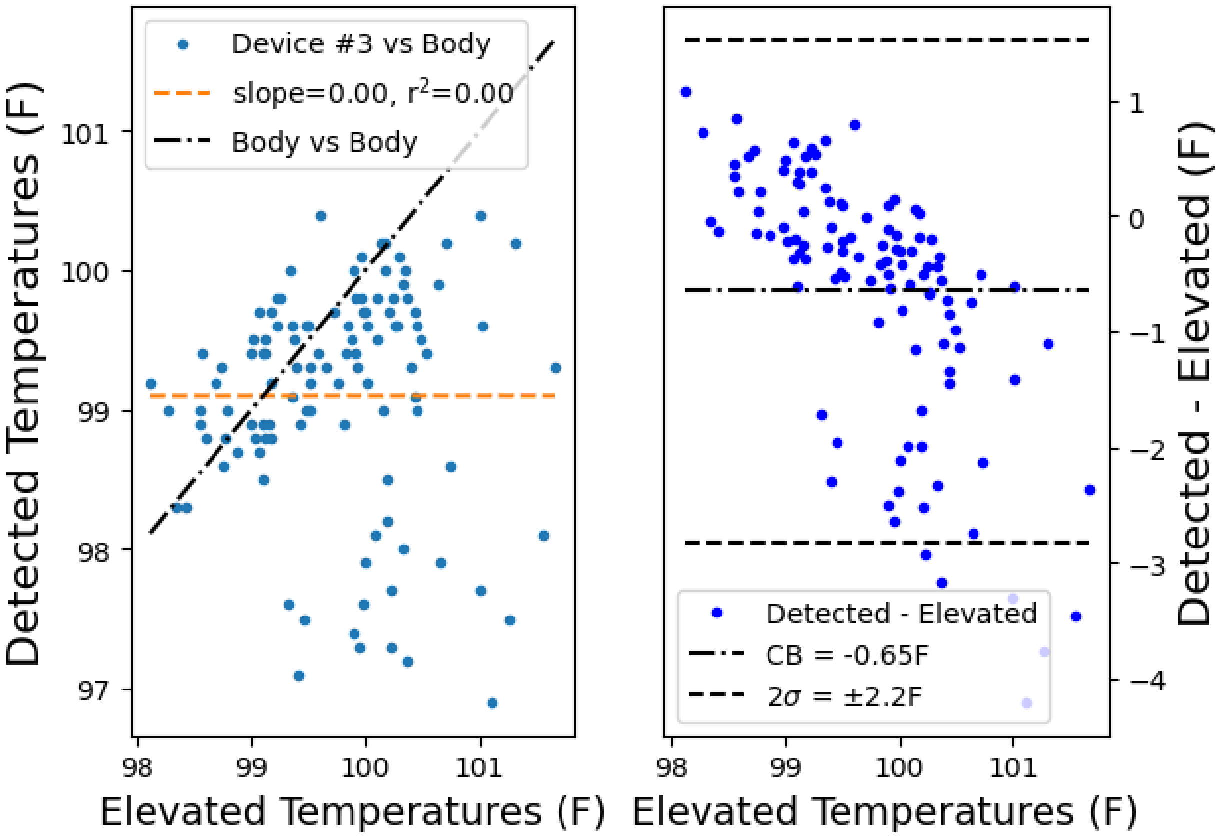

Device #3 vs Body
slope=0.00, $r^2$=0.00
Body vs Body
Detected Temperatures (F)
Elevated Temperatures (F)
Detected - Elevated
CB = -0.65F
$2\sigma$ = ±2.2F
Detected - Elevated (F)
Elevated Temperatures (F)

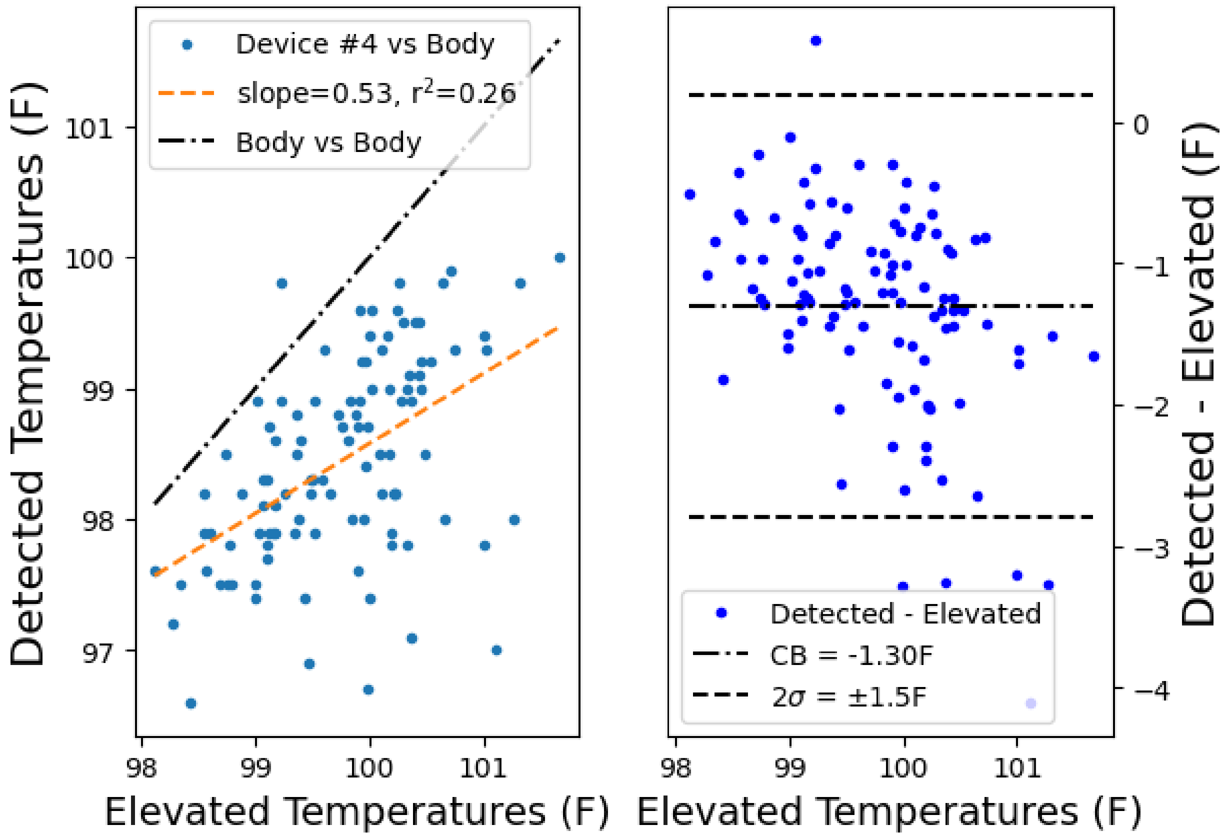
Device #4 vs Body
slope=0.53, $r^2$=0.26
Body vs Body
Detected Temperatures (F)
Elevated Temperatures (F)
101
100
99
98
97
98
99
100
101
Detected - Elevated
CB = -1.30F
2$\sigma$ = ±1.5F
Detected - Elevated (F)
Elevated Temperatures (F)
0
−1
−2
−3
−4
98
99
100
101

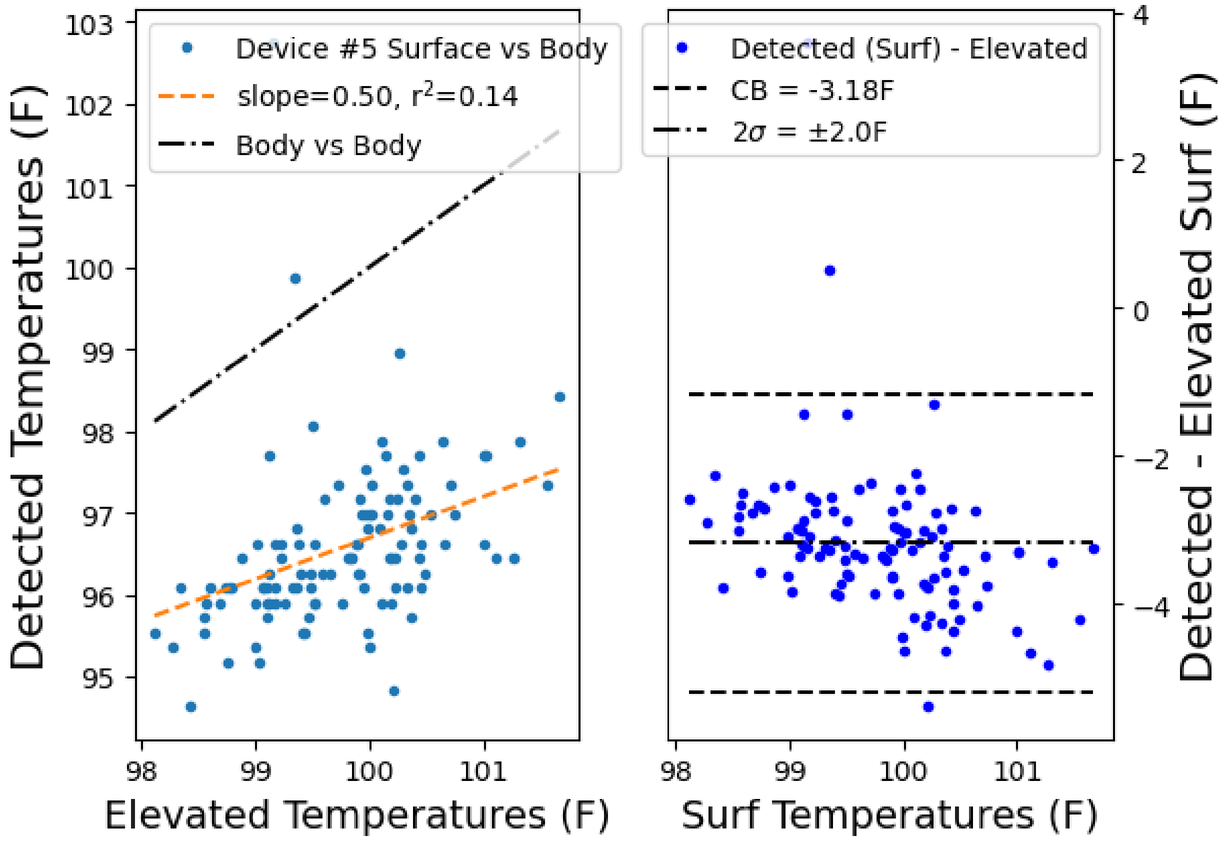


**Fig. S2** Left column shows detected vs simulated elevated temperatures and right column shows Bland-Altman plot (using elevated temperatures instead of the average in the x-axis due to potential bias in the detected temperatures), for each device. Note Device #3's slope and r2 are not typos as can be verified via the analysis script in the repository which explicitly prints these values to screen.

The clinical bias (CB) and limits of acceptability (LA, defined as $2\sigma$) are primarily dependent on the characteristics of the convenience sample of test subjects, and as such, could vary dramatically across test groups with different mean temperatures if there is any systematic bias, such as from a bias-to-normal algorithm. Regardless of the actual CB or LA value, a bigger concern is raised by the presence of a trend in the Bland-Altman plot, which can be caused by a bias-to-normal algorithm. Device #2 and Device #3 had prominent trend lines indicating bias-to-normal behavior, as seen in these plots. Thus, LA and CB are insufficient on their own to reveal this type of performance issue.

**Table S2** Linear fit slope coefficient for each device's detected body temperature (or surface, for Device #5) versus elevated body temperatures, with r-squared value for both elevated body temperature methods (prefixed with o for oral-based, s for surface-based).

| | oSlope | o $r^2$ | sSlope | s $r^2$ | oFPR | oTPR | sFPR | sTPR |
|---|---|---|---|---|---|---|---|---|
| Device 1 | 0.70 | 0.34 | 0.66 | 0.51 | 0.45 | 0.89 | 0.34 | 0.85 |
| Device 2 | 0.24 | 0.22 | 0.18 | 0.20 | 0 | 0.06 | 0 | 0.05 |
| Device 3 | 0 | 0 | 0.31 | 0.14 | 0.01 | 0.13 | 0 | 0.11 |
| Device 4 | 0.53 | 0.26 | 0.5 | 0.38 | 0 | 0 | 0 | 0 |
| Device 5 | 0.50 | 0.14 | 0.50 | 0.22 | 0.41 | 0.80 | 0.28 | 0.82 |

The false positive rates seen in Table S2 can be very large for Device #1, but note the design of this study results in an excess of borderline-threshold cases and as such, a large FPR is expected.

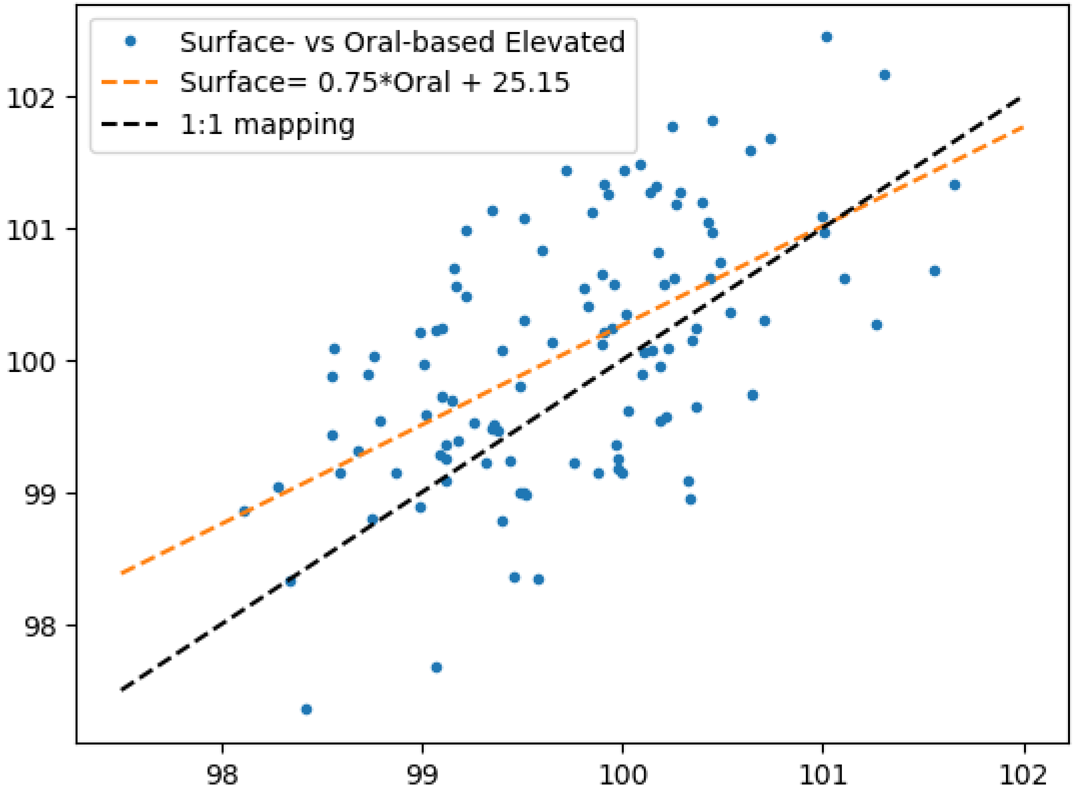


**Fig. S3** The two simulated elevated body calculation methods and relationship between them. The surface-based calculation produces values with approximately 25% lower spread.

*Radiative and Effective Temperature*

The effective temperature in each tent used for producing a simulated elevated body temperature was taken from the last 90 seconds of the measured air and radiative temperatures immediately prior to the acquisition by the five test devices. The difference between the radiative and air temperature in the tents was greater at higher air temperatures. In each case, the radiative temperature was monitored by capturing pixels of the background from each equilibration image and averaging these pixels. The effective temperature was produced by taking a weighted sum corresponding to the approximate balance between radiative and conductive air pathways, which was taken as 70% radiative, 30% conductive and are reported below in Table S3. The proportion of heat flow by the two pathways can be calculated but it must be pointed out there is some uncertainty in the literature regarding the heat transfer coefficient to air.

Under natural convection (with air speed under 0.5m/s), the heat transfer to air has been found to range from 3.1 to 5.1 W/(m$^2$K) **{49,50}**, while radiative can be calculated from the Stefan–Boltzmann law, using an emissivity of 0.98 (for both the muslin cloth radiative shielding used and the accepted value for human skin), a skin surface temperature of 34.5C (307.5K) and radiative temperature of 22C (295K), results in 76.1 W/m$^2$ for the radiative component and a range of 38.8 to 63.7W/m$^2$ for the non-radiative component. The fraction of each can be calculated for a fixed surface area, resulting in the radiative component comprising between 51 and 84% of the heat flow. As the radiative and air temperatures increase, the relative proportion of the radiative pathway will increase, but over the range used in this study the change is very small (less than 1%), and the dependence on ambient temperature can be approximated to within 1% by a linear relationship and hence Equations 1 and 2 can rely on a simple linear relationship instead of incorporating a 4th order polynomial arising from the radiative component. However, the range of heat transfer coefficients used here corresponds to averages over exposed tissues, which does not take into account the local geometry of body tissues. One study examined the effect of human body geometry on heat transfer **{51}** and found larger effects on heat transfer coefficients from more subtle changes in geometry and posture than can be appreciated in this study. Regarding the tissue region used in this study, the inner canthus is recessed by several millimeters, which would likely increase the thickness and resistivity of the local air boundary layer. For this reason, we increased the radiative proportion for inner canthi slightly to 70%. The overall effect on the effective temperature of reducing the radiative proportion from 70% to the lower theoretical fraction of 51% was examined by taking the average of the air and radiative temperatures of the warmest tent (Tent D), because this has the largest difference between air and radiative. The average Tent D air and radiative temperatures were 86.5F and 82.62F, with the effective temperatures for 70% radiative being 83.79F, while 51% radiative was 84.52F, a difference of 0.74F in effective temperature. Using Eqn 1, this corresponds to a potential systematic error of 0.12F in simulated elevated body temperature.

**Table S3** Temperatures for the last 90 seconds of air, radiative and combined effective temperature used in all analyses.

| Subject | TentA Air | TentB Air | TentC Air | TentD Air | TentA Rad | TentB Rad | TentC Rad | TentB Rad | TentA Eff | TentB Eff | TentC Eff | TentD Eff |
|---|---|---|---|---|---|---|---|---|---|---|---|---|
| E1 | 74.1 | 80.36 | 83.99 | 87.47 | 73.1 | 77.07 | 78.35 | 82.22 | 73.4 | 78.06 | 80.04 | 83.79 |
| E2 | 75.35 | 80.83 | 83.32 | 87.32 | 74.5 | 79.14 | 79.51 | 83.53 | 74.75 | 79.64 | 80.66 | 84.67 |
| E3 | 75.84 | 79.93 | 81.55 | 87.41 | 74.63 | 77.16 | 77.95 | 83.16 | 74.99 | 77.99 | 79.03 | 84.43 |
| E4 | 74.82 | 80.82 | 83.67 | 86.27 | 74.36 | 77.8 | 79.56 | 82.68 | 74.49 | 78.71 | 80.79 | 83.76 |
| F1 | 78.39 | 80.78 | 80.37 | 88.1 | 75.78 | 77.68 | 77.39 | 83.61 | 76.56 | 78.61 | 78.28 | 84.96 |
| F2 | 78.47 | 79.79 | 83 | 86.45 | 76.2 | 79.19 | 79.35 | 81.86 | 76.88 | 79.37 | 80.45 | 83.24 |
| F3 | 76.11 | 82.16 | 84.07 | 84.73 | 75.57 | 79.34 | 79.71 | 81.81 | 75.73 | 80.19 | 81.02 | 82.69 |
| F4 | 78.79 | 80.97 | 84.44 | 85.2 | 76.21 | 78.11 | 79.94 | 81.41 | 76.98 | 78.96 | 81.29 | 82.55 |
| F5 | 77.3 | 79.91 | 80.32 | 86.46 | 75.96 | 78 | 77.71 | 82.25 | 76.36 | 78.57 | 78.49 | 83.51 |
| F6 | 75.77 | 82.26 | 83.49 | 88.07 | 75.54 | 78.46 | 79.42 | 84.35 | 75.61 | 79.6 | 80.64 | 85.47 |
| F7 | 77.06 | 81.45 | 80.43 | 88.08 | 75.57 | 78.95 | 78.23 | 84.03 | 76.02 | 79.7 | 78.89 | 85.25 |

| | | | | | | | | | | | | |
|---|---|---|---|---|---|---|---|---|---|---|---|---|
| G1 | 76.9 | 78.27 | 81.6 | 86.23 | 78.04 | 79.2 | 79.63 | 83.99 | 77.69 | 78.92 | 80.22 | 84.66 |
| G2 | 77.4 | 82.72 | 83.13 | 84.93 | 75.61 | 80.28 | 79.64 | 82.37 | 76.15 | 81.01 | 80.69 | 83.14 |
| G3 | 76.98 | 81.98 | 84.46 | 87.24 | 75.94 | 79.37 | 80.37 | 83.54 | 76.26 | 80.15 | 81.6 | 84.65 |
| G4 | 78.6 | 80.74 | 84.68 | 85.5 | 76.01 | 77.76 | 79.58 | 81.65 | 76.79 | 78.65 | 81.11 | 82.81 |
| G5 | 76.29 | 82.15 | 83.62 | 86.47 | 75.56 | 79.62 | 79.48 | 83.37 | 75.78 | 80.38 | 80.72 | 84.3 |
| G6 | 77.51 | 81.03 | 83.47 | 87.24 | 75.71 | 78.59 | 78.95 | 82.92 | 76.25 | 79.33 | 80.31 | 84.22 |
| H1 | 75.95 | 80.17 | 81.96 | 85.41 | 75.01 | 78.58 | 79.38 | 82.4 | 75.29 | 79.06 | 80.15 | 83.31 |
| H2 | 76.84 | NA | 82.29 | 86.4 | 74.81 | NA | 79.91 | 82.27 | 75.42 | NA | 80.63 | 83.51 |
| H3 | 78.21 | 82.41 | 82.55 | 86.54 | 75.23 | 78.82 | 80.16 | 83.37 | 76.12 | 79.89 | 80.88 | 84.32 |
| H4 | 76.38 | 80.58 | 81.89 | 86.98 | 75.09 | 78.31 | 78.41 | 82.76 | 75.48 | 78.99 | 79.46 | 84.03 |
| I1 | 79.35 | 81.5 | 82.21 | 87.05 | 77.22 | 79 | 78.83 | 82.5 | 77.86 | 79.75 | 79.84 | 83.87 |
| I2 | 79.91 | 81.68 | 85.98 | 86.74 | 76.33 | 78.79 | 80.4 | 81.58 | 77.4 | 79.65 | 82.08 | 83.13 |
| I3 | 79.11 | 80.04 | 85.98 | 85.65 | 76.56 | 77.76 | 80.83 | 81.26 | 77.33 | 78.44 | 82.38 | 82.58 |
| J1 | 79.57 | 82.48 | 82.64 | 85.52 | 77.14 | 79.59 | 79.07 | 81.95 | 77.87 | 80.46 | 80.14 | 83.02 |
| J2 | 77.82 | 81.15 | 82.28 | 87.52 | 76.49 | 78.61 | 78.86 | 83.1 | 76.89 | 79.37 | 79.89 | 84.42 |
| J3 | 78.97 | 80.01 | 82.96 | 85.78 | 76.91 | 77.86 | 78.81 | 81.71 | 77.53 | 78.51 | 80.05 | 82.93 |
| J4 | 78.83 | 81.15 | 85.72 | 85.33 | 76.56 | 78.56 | 80.72 | 81.79 | 77.24 | 79.34 | 82.22 | 82.85 |

**Table S4** Temperature measured by each device after the warmest equilibration period (Tent D).

| Subject | Device 1 | Device 2 | Device 3 | Device 4 | Device 5 |
|---|---|---|---|---|---|
| E1 | 99.87 | 98.4 | 99.7 | 98.2 | 94.82 |
| E2 | 101.24 | 98.5 | 99.0 | 99.2 | 96.08 |
| E3 | 99.74 | 98.1 | 96.9 | 97.0 | 96.44 |
| E4 | 101.19 | 98.3 | 97.7 | 97.8 | 96.62 |
| F1 | 101.73 | 98.5 | 99.6 | 98.9 | 96.62 |
| F2 | 102.04 | 98.9 | 99.8 | 99.6 | 97.16 |
| F3 | 101.91 | 98.5 | 100.1 | 99.5 | 97.52 |
| F4 | 101.43 | 98.5 | 98.1 | NA | 97.34 |
| F5 | 100.96 | 98.5 | 99.6 | 98.9 | 96.44 |
| F6 | 101.56 | 98.7 | 98.6 | 99.3 | 96.98 |
| F7 | 101.60 | 98.8 | 100.2 | 99.8 | 97.88 |
| G1 | 101.05 | 98.5 | 99.1 | 99.1 | 96.44 |
| G2 | 100.25 | 98.5 | 97.9 | 98.0 | 96.62 |
| G3 | 100.65 | 98.4 | 100.1 | 98.4 | 96.98 |
| G4 | 100.66 | 98.6 | 99.9 | 99.0 | 97.34 |
| G5 | 101.12 | 98.3 | 99.1 | 98.8 | 96.80 |
| G6 | 100.36 | 98.5 | 100.0 | 98.7 | 96.62 |
| H1 | 10051 | 98.7 | 99.6 | 99.0 | 96.62 |
| H2 | 101.13 | 98.5 | 99.6 | 99.8 | 98.96 |

| | | | | | |
|---|---|---|---|---|---|
| H3 | 102.48 | 99.4 | 100.4 | 99.4 | 97.70 |
| H4 | 102.56 | 99.8 | 99.3 | 100.0 | 98.82 |
| I1 | 100.48 | 98.4 | 99.6 | 98.0 | 96.44 |
| I2 | 100.63 | 98.5 | 99.8 | 98.9 | 96.80 |
| I3 | 101.71 | 99.5 | 100.2 | 99.9 | 97.34 |
| J1 | 101.13 | 98.8 | 99.8 | 99.3 | 97.88 |
| J2 | 100.76 | 98.4 | 97.5 | 98.0 | 96.44 |
| J3 | 100.51 | 98.5 | 100.0 | 99.1 | 96.98 |
| J4 | 99.52 | 98.4 | 99.0 | 97.9 | 95.9 |

*Manufacturer Specifications*

**Table S5** Manufacturer device specifications and usage.

| Device | Minimum Pixel Size | Operating Distance | Ambient Correction | DE/SSE Correction | Array Size | Method |
|---|---|---|---|---|---|---|
| 1 | 1.5mm | 0.3 - 1.6m | Yes | Yes/Yes | 80x80 | Canthi |
| 2 | 1.4mm | 0.5 - ? | No | No | 160x120 | Face |
| 3 | NA | 5mm | No | NA | 1 | Sweep |
| 4 | 4.4mm | 1.52m | No | NA | 206x156 | Face |
| 5 | 1.6mm | 1m - ? | No | No | 464x348 | Face |

The specifications of each device are given in Table S5, with a smallest pixel size based on the field of view, array size and shortest/recommended distance to subject for each device. Note, Device 1 has a large range up to 1.4 meters, which results in a pixel spot size of 8.0mm (23° FOV). Device 2 and Device 5 do not specify a shortest range but based on the user interface, it may be challenging to obtain routine measurements from shorter than 1 meter distance. SSE and DE was only measured for Devices 1 and 5.

Device 1 uses a single-shot convolutional neural network face detector trained on and applied to thermal images to identify a crop covering from the eyebrows to the center of the nose and laterally covering to the outer corners of both eyes and takes the median of the hottest 8 pixels inside this region, after linearizing the full image to two blackbodies in the field of view maintained at 32 and 37C and applying an SSE correction. The resulting temperature is then corrected with an empirical DE correction for inner canthi to obtain a distance-corrected surface temperature which is used in the present study as the equilibration surface temperatures and this corrected surface temperature is finally corrected for air temperature to obtain a body estimate. Device 2 is an industrial IRT with an improved calibration accuracy for body ranges and reports the hottest pixel within the full field of view. Device 3 consists of a plastic cup-shaped probe holding a single-pixel thermopile sensor recessed approximately 5mm from its opening, which is intended to be swept while gently pressing against the subject's forehead from midline to temple while pressing a button, releasing said button when the sweep is complete, whereupon the device reports some adjusted body temperature based on the readings sampled during the sweep. Device 4 uses a face detector operating on a paired visible-light (color) USB camera with the detected face bounding box transformed to the thermal camera coordinates and the detected maximum is offset-adjusted by a resistively heated blackbody in the field of view. Device 5 displays a head outline on the display screen overlaid on the continuously updating thermal image stream, and the intended use of the device is for the operator of the device to ensure the subject's frontal face view is contained within this head outline (note, different sized head outlines can be selected in the configuration options in order to configure the outline to best match the size of the average human head at different distances to the subject) and the maximum temperature pixel is identified and compared to a moving average of the last 8 measurements. Note, the specific details of the processing that occurs in Devices 2 through 5 may differ slightly from these descriptions.

*Bias-to-normal in quick-read mode oral thermometry*

The oral thermometer used in this study (Welch Allyn SureTemp Plus 690) operates in two modes: quick-read mode, which returns an oral temperature within approximately 10 seconds and has a specified clinical accuracy of 0.2C, and monitor mode, which simply reports the evolving temperature measured at the probe tip and which has a specified clinical accuracy of 0.1C. The quick-read mode fits an exponential decay to calculate the expected final temperature, but if the time constant is comparable to the measurement time, any variability in those measurements will produce unacceptably large errors in the projected final temperature. Based on the time required for readings returned in the device's monitor mode, which was up to 120 seconds to stabilize within 0.1C, the time consta-nt is likely between 10 and 30 seconds. Changing the first or last value by 0.1C in 10 exponentially decaying measurements over 10 seconds with a time constant of 15 seconds results in a projection error as large as 0.6C, which is comparable to what was observed in several of the subjects having oral temperatures further from 98.6F in Table 3. Fitting the quick-read versus monitor mode readings in Fig. S4 shows what may be a bias-to-normal implemented in order to eliminate larger misprojection errors from the exponential fit process. Regardless, this device is frequently used in studies of human body temperature, and designers of body temperature studies should be aware that what is specified as a 0.2C accuracy for quick-read mode may be misleading when measuring temperatures that differ from the population average.

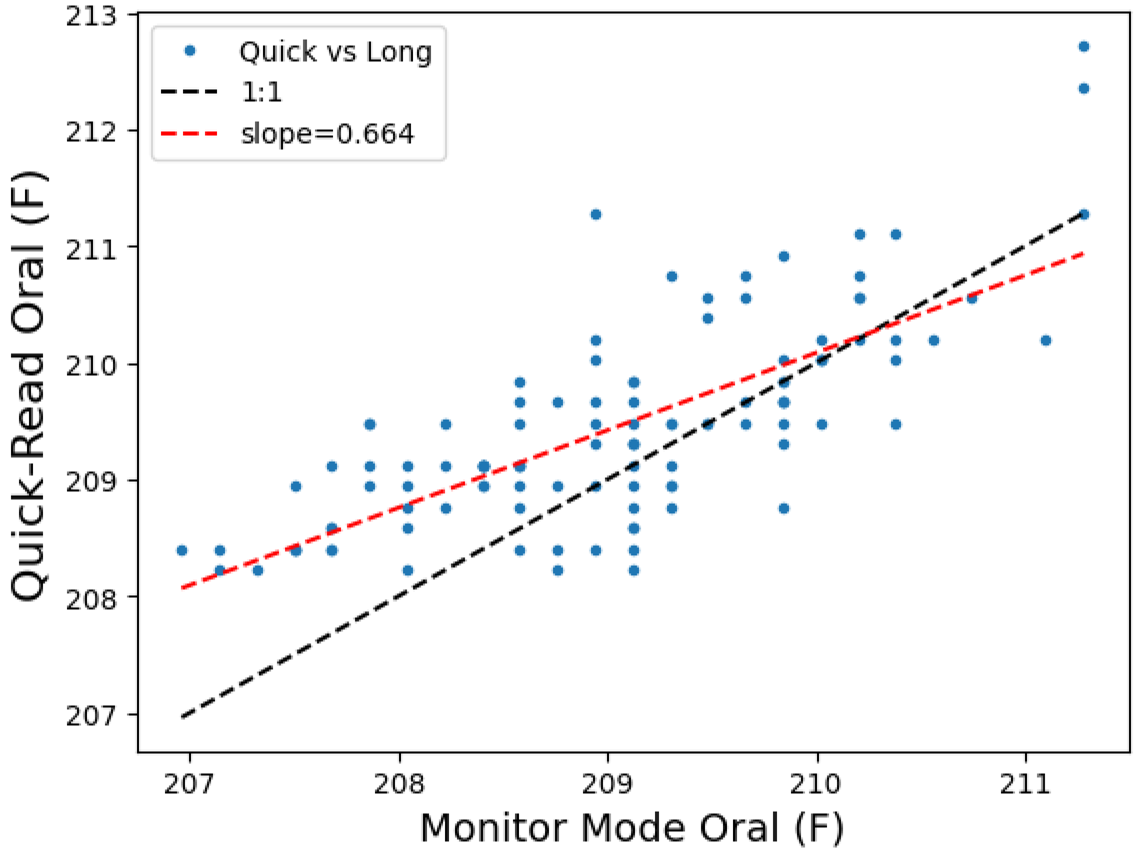


**Fig. S4** Quick read (y-axis) versus monitor mode oral thermometry readings from Table 3 and linear fit between the two mode readings.
